# Benchmarking autosomal recessive disease prevalence estimation from allele frequencies against newborn screening data

**DOI:** 10.1101/2025.10.11.25337773

**Authors:** Michael C Sierant, Nick Knoblauch, Evan Witt, Daniel Gaffney, Sara L Pulit, Arthur Wuster

## Abstract

Accurate estimates for the prevalence of rare congenital diseases are critical for understanding disease epidemiology and enabling drug development. Prevalence estimates can inform public health investment, identify communities with high disease burden or underdiagnosis, and reveal areas of unmet clinical need.

With the advent of global-scale biobanks, genetics-based models to estimate the prevalence of disease have become viable. Autosomal recessive (AR) rare diseases are particularly tractable for this approach given that disease prevalence can be estimated from the pathogenic allele frequency (AF) in carriers from unaffected populations. Despite the usefulness of such estimates, this approach has not been validated against real-world clinical datasets at scale. Newborn screening (NBS) programs, which test newborns for a panel of neonatal diseases using quantitative diagnostic methods, provide a comparator for birth prevalence with low ascertainment bias, large sample size, and low diagnostic variability. NBS datasets thus offer a uniquely robust benchmark to evaluate and improve the accuracy of AR genetic prevalence models.

Here we explore the feasibility, utility, and pitfalls of estimating AR birth prevalence using genetic and NBS data. We applied a genetic model to estimate birth prevalence for 28 AR diseases consistently present on NBS panels and benchmarked these against reported NBS birth prevalence in more than 12 million newborns in the United States. We found concordance between the genetic estimate and NBS was impacted by the population database used to derive AF, ancestry-matching methodology, and pathogenic variant inclusion criteria. Incorporating these refinements, we demonstrate that a genetics-first approach can provide order-of-magnitude estimates of AR disease birth prevalence for nearly all tested diseases (25/28; 89%). However, we note a general underestimate of the genetic prevalence, suggesting identifying additional pathogenic variants would improve the concordance with NBS. Further, we also assessed the impact of epidemiological and genetic variables, highlighting diseases where genetic prevalence estimates may not be suitable.

## Introduction

Accurately estimating the birth prevalence of rare congenital diseases, which affect approximately 300 million people worldwide,^1^ is a fundamental component of understanding disease epidemiology. High-quality prevalence estimates can inform public health investment, identify communities with high disease burden or underdiagnosis, and reveal areas of unmet need. Additionally, drug discovery and development rely on prevalence estimates to assess patient populations and prioritize clinical programs.

Multiple approaches exist for assessing disease prevalence. Most commonly, epidemiology studies estimate prevalence by counting diagnosed patients using hospital records or performing systematic screening in a defined region. Although this approach directly counts diagnosed patients, diseases that are difficult to clinically assess due to complex phenotypic presentations may be underdiagnosed in a population^2^ and therefore underestimated.

An alternative approach is newborn screening (NBS). Heel prick samples are drawn from newborns, and the samples are run through a battery of quantitative biochemical diagnostic methods. In the U.S., neonates are tested for approximately 30 early-onset disorders as a public health initiative.^3^ Unlike traditional epidemiological efforts, quantitative testing of well-established biomarkers of disease (for example, biochemical testing for high phenylalanine levels in phenylketonuria^4^) allows for diagnosis prior to onset of disease. Critically, NBS in the U.S. has near-universal uptake (>99% of newborns) which minimizes ascertainment bias.^5,6^ However, inconsistent nationwide implementation of testing methodologies or patchwork inclusion of diseases (for example, the NBS panel is not standardized state to state) can contribute to imprecise estimates.^3,7,8^ Further, existing tests for some diseases do not have perfect predictive power, resulting in an elevated false negative rate (for example, patients with intermediate galactocerebrosidase activity can still develop Krabbe disease).^9^ Most problematically, implementation of NBS for new diseases at nationwide scale is resource intensive and slow with a submission-to-approval process that takes on average 9 years.^10,11^ Consequently, U.S. NBS programs are limited to fewer than three dozen to date, while other nations sometimes adopt fewer.^12–15^ Nonetheless, NBS data is likely the most accurate source for birth prevalence currently available.^13,16–18^

More recently, genetic data has been used to estimate the number of individuals harboring pathogenic genotypes and thereby predict the prevalence of disease phenotypes.^19,20^ This approach uses allele frequencies in large population cohorts such as the UK Biobank^21^ or gnomAD^22^, is scalable, and can result in accurate estimates;^23–26^ but, challenges can include undiscovered or misannotated disease-causing mutations, variable penetrance of mutations^7^, violations of Hardy-Weinberg equilibrium due to nonrandom mating, and imprecision of AF measurements for rare mutations - especially in individuals of non-European descent who are poorly represented in biobanks.^29^ Orthogonal to traditional epidemiological approaches, genetic methods offer a patient-free method to understand birth prevalence for a wider array of diseases at scale, especially extremely rare diseases that are difficult to quantify without extremely large sample sizes.

Given the widespread availability of genomic data, the genetic approach to estimating AR disease birth prevalence has become increasingly common;^20,23–25,30–32^ however, its accuracy has not been systematically assessed by comparing directly to available NBS data. We therefore sought to generate birth prevalence estimates from genetic data (hereafter designated as BP_g_) and compare them to birth prevalence derived from NBS data (BP_NBS_). Specifically, we used publicly available allele frequencies from gnomAD to estimate the birth prevalence of 28 AR diseases that are screened for in newborns across all 50 states. We generated baseline estimates using variants with clinical evidence for a role in disease and then compared these to the NBS data. We then iteratively explored the impact of allele frequency database, representation of ancestral groups, and inclusion of additional tiers of variant pathogenicity in generating birth prevalence estimates. Across our analyses, we found that genetics can provide reliable estimates of AR disease birth prevalence across NBS diseases with limited sources of genetic and epidemiological input. However, we note a general underestimate of the prevalence compared to NBS and explore when caution is necessary when interpreting reported prevalence.

## Methods

### Selection of newborn screening (NBS) datasets

As of 2020, approximately 35 diseases are included on the Recommended Uniform Screening Panel (RUSP) for which newborns are tested in the United States (U.S.).^33^ This system is implemented state by state, a feature that complicates data aggregation but has some advantages. For example, some states choose to collect additional demographic data beyond RUSP recommendations – one example being California’s collection of self-reported newborn ethnicity, data that enables both pooled and ethnicity-specific analyses.^8^ To benchmark disease birth prevalence, we used two data sources: (1) NBS data aggregated across the U.S., Puerto Rico, and Guam for 2015-2020 which totals ∼22M screened individuals (hereafter designated as ‘nationwide NBS’)^33^ and (2) NBS data for California for 2009-2019 which totals ∼5.35M screened individuals and includes self-reported ancestry (hereafter designated as ‘California NBS’).^34^

In the nationwide data, most diseases had been screened for in ∼11M births, with the exception of diseases that the Advisory Committee on Heritable Disorders in Newborns and Children has only more recently recommended be added to the RUSP: Pompe disease (∼5.5M screens, added in 2015^35^), Mucopolysaccharidosis I (∼5.1M screens, added in 2016^36^), and spinal muscular atrophy (∼3.2M screens, added in 2018^37^). Concordantly, the total individuals screened for these diseases is lower in the California data. Finally, the RUSP is a recommendation only and some states choose not to screen for certain diseases; these diseases are excluded entirely due to state-specific data reporting or NBS inclusion criteria.

For the benchmarking process, we restricted our analyses to the 28 autosomal recessive disorders included in our selected NBS panels (**Table 1**). We calculated birth prevalence for our diseases of interest per 100,000 people by first dividing the number of reported positive screens (cases) by the total number of screens and then multiplying by 100,000. Ninety-five percent confidence intervals were calculated using the Clopper-Pearson method.

**Table 1.**
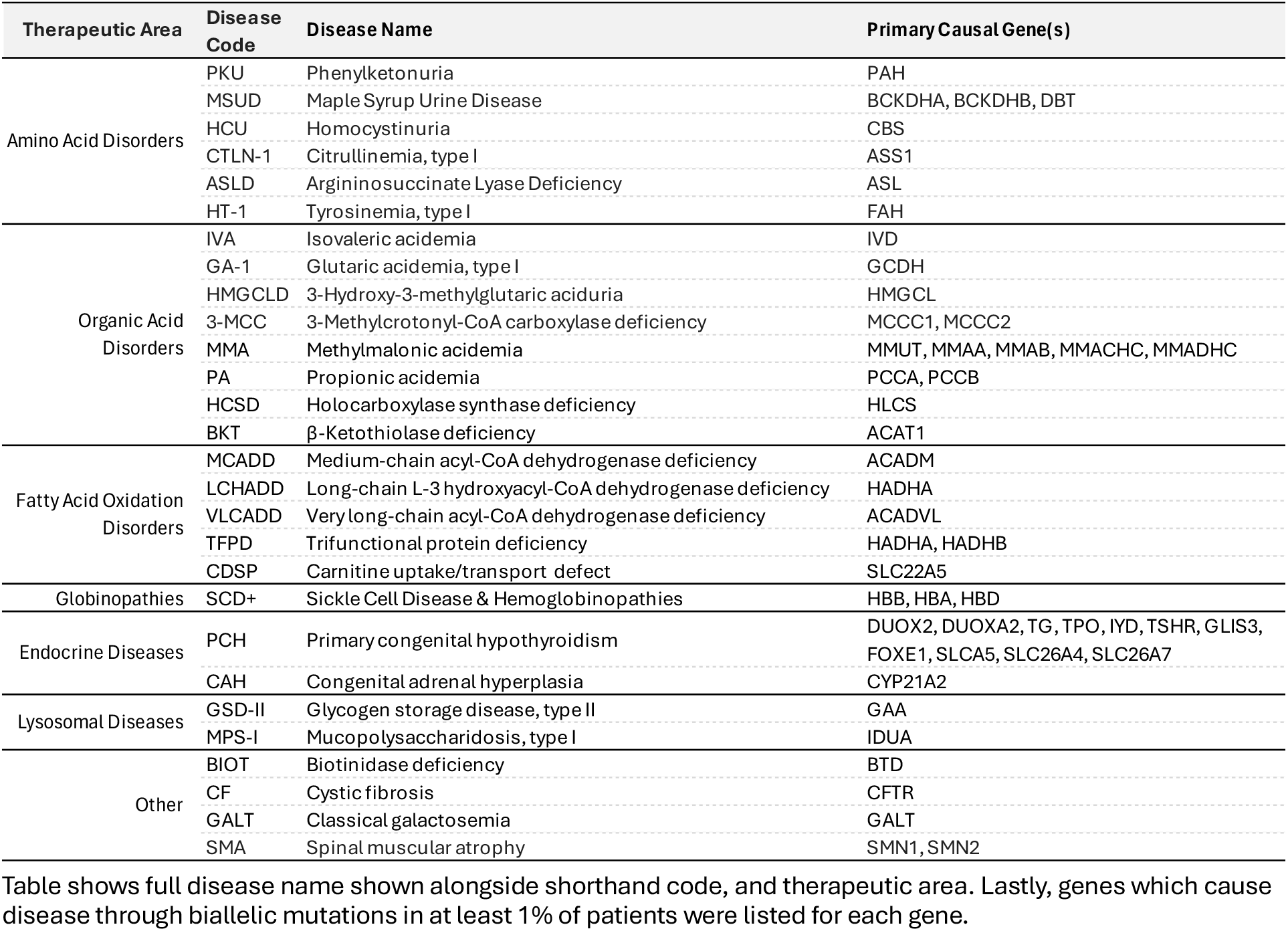
Twenty-eight selected autosomal recessive NBS diseases for prevalence benchmarking.

### Calculating birth prevalence of autosomal recessive (AR) disease

To generate genetics-based birth prevalence estimates, we used variant allele frequencies reported in gnomAD version 2.1.1^38^, which contains exome and genome data from 141,456 individuals (125,748 exomes and 15,708 genomes); and version 4.1,^22^ which contains data from 807,162 individuals (730,947 exomes and 76,215 genomes^22^) aggregated across >9 ancestral populations.

We analysed allele frequencies from gnomAD release version 2.1.1 to investigate the impact of using allele frequencies from a biobank with fewer samples (compared to e.g., gnomAD v4.1). Because v2.1.1 was released on genome build GRCh37, we used the lifted over data from GRCh37 to the GRCh38 reference genome.

To estimate disease birth prevalence, we assumed that all mutations included in our calculations were fully penetrant and that their presence in either a homozygous or compound heterozygous state in an individual resulted in disease. We did not account for linkage disequilibrium in our estimates; pathogenic variants tend to be rare (and therefore independent of one another) due to selective pressure.

We generated variant-based birth prevalences estimates for the whole gnomAD population, and then separately for each subpopulation. We compared these estimates to the California NBS birth prevalences from the whole population as well as within self-identified ethnicity groups. We visualized these comparisons with ggplot2 and made statistical comparisons with `ggpubr::stat_cor()`.

### Derivation of the prevalence formula

We consider a scenario with multiple independent loci, each with a recessive disease-causing allele that can contribute to the same condition. Let *p* represent the number of such loci, and *f_i_* be the frequency of the disease-causing allele at locus *i*. Under Hardy-Weinberg equilibrium (HWE), the probability that an individual inherits the disease allele from a given parent at a specific locus is simply *f_i_*, and conversely, 1-*f_i_* for not inheriting the disease allele. Given *p* independent biallelic loci, each with a disease-causing allele at frequency *f_i_*, the probability of a single copy of a gene *not* containing a disease allele at any locus (i.e., the probability of having the wild type allele at every locus in the gene) is 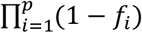. The complement of this probability, 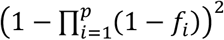, represents the probability that an individual inherits at least one disease-causing allele.

To find the probability of an individual being affected (inheriting at least one disease-causing allele on each of two copies of a gene), we consider the probability of a parent not transmitting any disease allele, 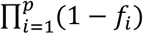. The complement, 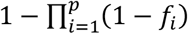, is the probability of a parent transmitting at least one disease-causing allele. Squaring this value to yield 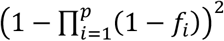 gives the probability of both parents transmitting at least one disease-causing allele or, equivalently, the probability that the parents’ offspring will be affected by disease. Thus, the estimated birth prevalence per 100,000 births is 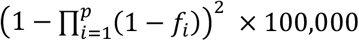.

### Extension to populations with multiple ancestries

The birth prevalence derivation above assumes a single, homogenous population. However, real populations often consist of individuals from diverse ancestral backgrounds. When allele frequencies vary significantly between these groups, it becomes necessary to account for this population structure. Here, we extend our model to a population composed of *k* distinct ancestry groups, ignoring the possibility of admixture between these groups.

Let w_j_ represent the proportion of individuals in the overall population belonging to ancestry group *j*, such that 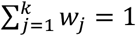. Let f_i,j_ be the frequency of the disease-causing allele at locus *i* in ancestry group *j*. To obtain the overall population prevalence, we take a weighted average of the prevalences in each ancestry group, weighted by their respective proportions in the population, so 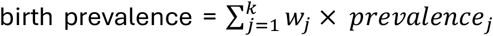. Substituting the expression for *prevalencej* (above) we get:

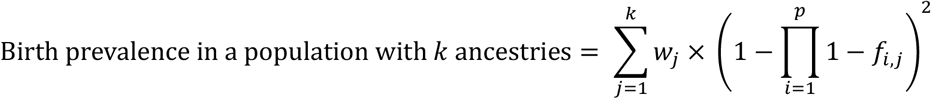

### Comparing prevalence between versions of gnomAD for individual genes

To compare the birth prevalence of a recessive genetic trait between two cohorts (e.g., between gnomAD v2.1.1 and gnomAD v4.1), we extended this prevalence estimation framework to perform hypothesis testing using the Wald test, formalized here.

The variance of 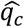 (the recessive prevalence in cohort 𝑐) is approximated using the delta method, which accounts for the non-linear transformation 𝑞 = (1 − ∏_𝑖_ (1 − 𝑓_𝑖_))^2^. Let 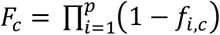 represent the probability that a single chromosome carries no recessive alleles at any locus in the cohort (*c*). The variance of 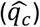 is 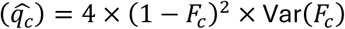, where Var(𝐹_𝑐_) is derived using the delta method on F_c_:

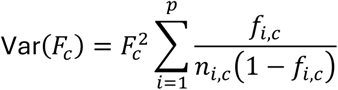

Here, 𝑛_𝑖,𝑐_ is the total number of alleles observed at locus 𝑖 in cohort *c*. Substituting into 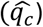, we obtain:

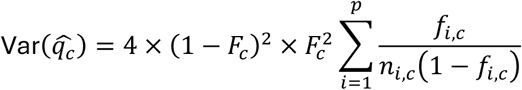

To test the null hypothesis (H_0_) that the prevalences are the same (q_1_ = q_2_), we compute the Z-score for the difference in prevalence estimates:

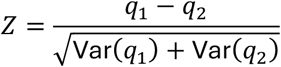

Under 𝐻_0_, 𝑍 asymptotically follows a standard normal distribution. The two-tailed p-value is calculated as p = 2 × 𝛷(−|𝑍|) where (𝛷) is the cumulative distribution function of the standard normal distribution.

### Variant effect prediction

Variants in the gnomAD database are already annotated with pathogenicity classifications from ClinVar. Variant pathogenicity classifications were based on the latest release of ClinVar^39^ (updated August 2024). For birth prevalence estimates based on ClinVar annotations only, we selected the set of variants annotated as “pathogenic,” “likely pathogenic,” or “pathogenic/likely pathogenic,” and implicated in the specific disease we were studying.

We also annotated variants using *in silico* prediction tools. gnomAD data comes with loss-of-function variant annotation generated using LOFTEE^38^; we excluded loss-of-function variants that failed the LOFTEE filtering criteria (non-PASS). Additionally, we annotated variants using precomputed variant-specific scores from the AlphaMissense^40^ and REVEL^41^ plugins included in the Ensembl Variant Effect Predictor^42^ (VEP) tool. Missense variant counts from gnomAD and annotations from ClinVar were merged with variant effect predictions from a modified version of the Ensembl VEP pipeline with the REVEL plugin.

### Calculation of gene coverage in gnomAD exome and genome data

To determine how well a gene is captured in a given population database (*e.g.* gnomAD v2 or v4), we calculated a dropout metric of base-sample pairs that accounts for the number of samples across all assessed bases with fewer than 10 aligned reads. For a given gene, we defined this region as the coding exons plus 10bp of flanking region to include potential splice-disrupting variants. The dropout metric was calculated as:

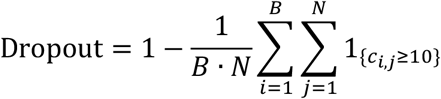

Here: B is the total number of targeted bases, N is the total number of samples, and C_i,j_ is the coverage at base *i* in sample *j*.

### Comparison between newborn screening and genetic prevalence

Concordance between log10-transformed BP_g_ versus BP_NBS_ was quantified using Lin’s Concordance Correlation Coefficient (ρ_c_).^43^ We apply ρ_c_ here, rather than use Pearson’s correlation, because it evaluates both precision and accuracy by penalizing systematic differences between measurements to determine deviation from perfect agreement, rather than just assessing for the strength of a linear relationship.

### U.S. Census Data

We downloaded the latest census data available through 2024 for all residents of the United States (U.S.) (called ‘Universe R’ by the U.S. Census Bureau^44^). Data was reported for the whole population as well as stratified by self-identified ethnicity.^45^

### Mapping genetic ancestry from gnomAD to self-identified ethnicity groups

To explore the impacts of genetic ancestry on our birth prevalence estimates, we made comparisons between self-identified ethnicity groups in the California NBS and census data to ancestry groups in gnomAD participants that were quantitatively classified using genetic data.

We matched genetic ancestral groups in gnomAD to self-identified ethnicity groups in the census data as others have done^22,38^, relying on additional demographic information to broadly align groups. To accomplish this, we were required to make a few assumptions about the US census and CA NBS self-report data. First: people of Middle Eastern and North African descent are counted as “white” in the U.S. census.^46^ Currently, people who identify as Middle Eastern or North African alone account for approximately 1.1% of the U.S. population.^47^ We therefore assumed that 1.1% of the “white alone” group in the census data matched to people of Middle Eastern descent in gnomAD. Second: there are approximately 5.8 million Jewish adults living in the United States (U.S.), and another 1.8 million children living with at least one Jewish adult.^48^ Approximately two thirds of Jewish people in the U.S. are Ashkenazi Jewish^21^ and we have assumed that all would identify as “white alone” when self-identifying. Thus, we assumed that approximately 1.5% of the U.S. and CA population would map to the Ashkenazi Jewish ancestral group in gnomAD. Third, exclusive to the census data: although individuals can identify as “Asian” on the U.S. census, they cannot identify with a subregion of the Asian diaspora. Approximately 6.5 million people in the U.S. are South Asian Americans^49^ and we have mapped these individuals to the South Asian ancestry group in gnomAD. We have assumed that the remaining individuals in the “Asian” census category can be mapped to individuals of East Asian descent in gnomAD. Fourth, exclusive to the CA NBS data: individuals reported as “Southeast Asian” or “Pacific Islander” were mapped onto the East Asian group in gnomAD, as these groups strongly genetically cluster to those of East Asian ancestry.^50,51^ Fifth: those self-identified as ‘Hispanic’, ‘Latino’, or ‘Native American’, or ‘American Indian’ were matched to the ‘Admixed American’ group in gnomAD.

## Results

### A simple genetic model underestimates NBS prevalence

We estimated the birth prevalence of 28 AR diseases, which we refer to using their abbreviations (**Table 1**), using genetic methods and then compared them to prevalence estimated from NBS datasets (**Methods**). We use the term *birth prevalence* (BP) throughout to describe our findings and in accordance with standard epidemiological practice.^52^ Unless otherwise stated, the unit of BP is the number of affected individuals per 100,000 live births.

For our initial baseline analysis, we used minimal prior knowledge about the genetic or epidemiological architecture of the disease to improve the generalizability of the approach and understand the importance of each additional component. We extracted the aggregate AFs for pathogenic mutations across all participants from the most recent version of the gnomAD database (v4.1). The definition used for pathogenic variants in this baseline model were those labeled as pathogenic or likely pathogenic (P/LP) in the ClinVar database.^39,53^ From this, we calculated the predicted birth prevalence of individuals with a biallelic genotype in the gnomAD sample population (**Methods**).

We then compared our genetics-derived estimate (BP_g_) against the birth prevalence of each disease reported from NBS (BP_NBS_) using the U.S. nationwide dataset. To measure concordance, we used Lin’s concordance correlation coefficient (ρ_c_) which, in addition to assessing the strength of the linear relationship, also captures any deviation of BP_g_ from perfect agreement with BP_NBS_.^54^ Our initial comparisons revealed moderate concordance (Lin’s ρ_c_ = 0.61 [0.38-0.76 95% CI]) with 5/28 (18%) of diseases within 2x of the genetic estimate and 17/28 (61%) within 10x (**Figure 1A**). We found that BP_g_ was systematically lower than BP_NBS_ by a median of 3.4-fold across all diseases. For 10/28 diseases (36%), BP_g_ was 10% or less of the prevalence reported by NBS. Six diseases were notable outliers, deviating by more than 30x from BP_NBS_: HCSD (BP_g_ 78x lower than BP_NBS_), PA (60x lower), 3-MCC (36x lower), HMGCLD (30x lower), and SMA (74x higher); the only overestimate was BIOT (32x higher) (**Figure 1C**) (**Table 2**). We explored whether simple modifications to this approach could produce better concordance between BP_g_ and BP_NBS_.

**Figure 1.**
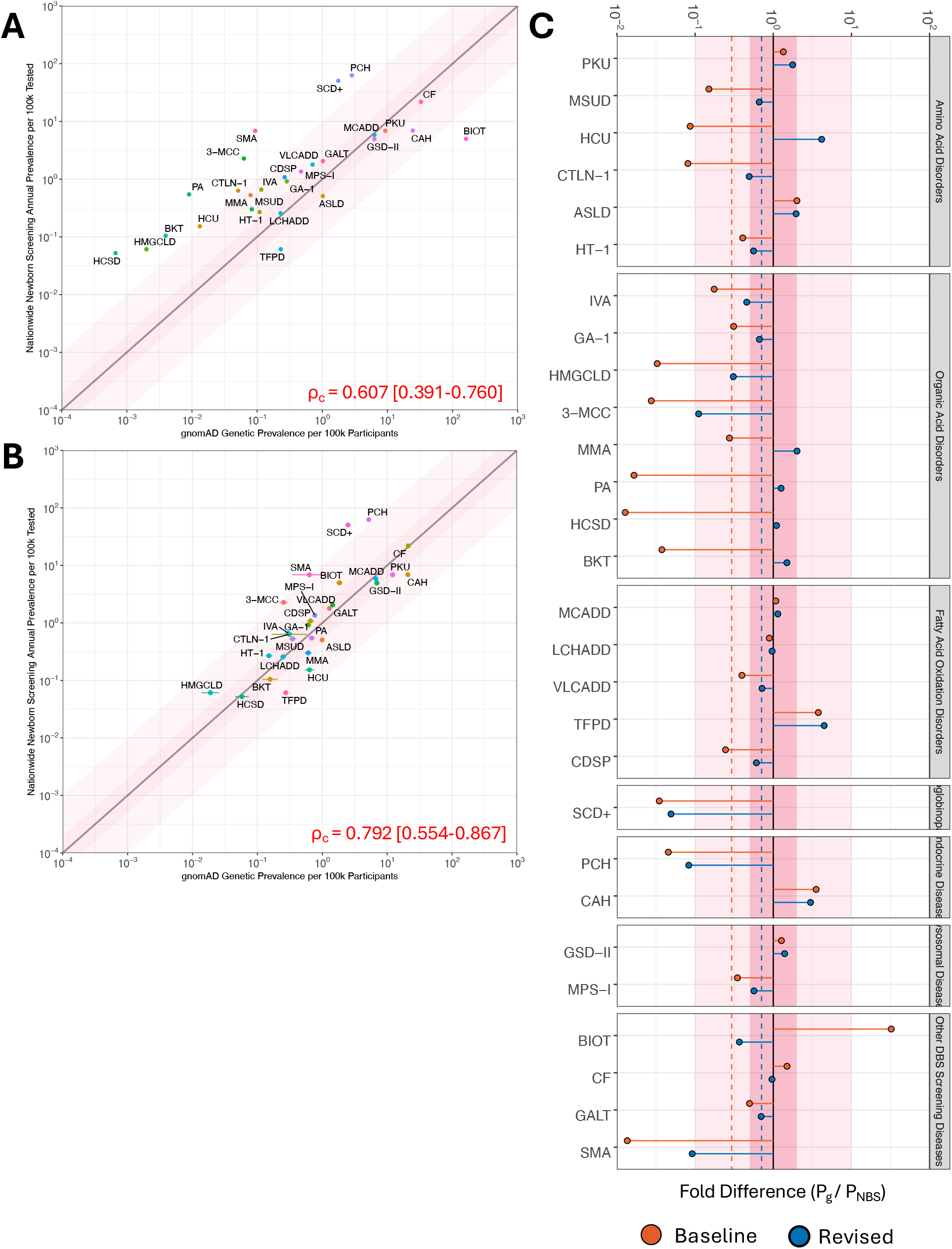
Concordance between genetic prevalence of AR disease and empirical NBS. **[A]** Scatterplot of baseline genetic estimated prevalence (x-axis) versus the reported nationwide NBS prevalence (y-axis) for 28 AR diseases, both shown per 100k. The line of equality is shown in black with 2x and 10x ribbons in dark pink and pink, respectively. Lin’s concordance correlation coefficient (ρ_c_) is annotated in red. **[B]** Same as [A], but using revised genetic prevalence estimation. **[C]** Lollipop plot of the fold-difference in prevalence for each disease between the genetic estimate and reported NBS prevalence. Results from the baseline genetic model are shown in orange and revised in blue. Median FD across all diseases are shown as dashed lines and annotated for each model, excluding HCU in the calculation. The 2x and 10x FD ranges are shown in dark pink and pink, respectively. Diseases are stratified by therapeutic area per Table 1.

**Table 2.** Concordance between genetic estimate and NBS for AR diseases using initial and revised models.

| Therapeutic Area | Disease Code | NBS Prevalence |  |  | Genetic Prevalence |  |  |  |
| --- | --- | --- | --- | --- | --- | --- | --- | --- |
|  |  | Cases | Births | :100k | Baseline |  | Revised |  |
|  |  |  |  |  | :100k | Ratio | :100k | Ratio |
| Amino Acid Disorders | PKU | 1,576 | 22,930,291 | 6.873 | 9.256 | 1.35 | 12.168 | 1.77 |
|  | MSUD | 121 | 22,930,291 | 0.528 | 0.079 | -6.66 | 0.349 | -1.51 |
|  | HCU | 35 | 22,930,291 | 0.153 | 0.013 | -11.56 | 0.635 | 4.16 |
|  | CTLN-1 | 145 | 22,930,291 | 0.632 | 0.051 | -12.36 | 0.311 | -2.03 |
|  | ASLD | 116 | 22,839,659 | 0.508 | 1.020 | 2.01 | 0.996 | 1.96 |
|  | HT-1 | 61 | 22,695,225 | 0.269 | 0.109 | -2.46 | 0.150 | -1.79 |
| Organic Acid Disorders | IVA | 151 | 22,930,291 | 0.659 | 0.116 | -5.70 | 0.301 | -2.19 |
|  | GA-1 | 210 | 22,930,291 | 0.916 | 0.285 | -3.21 | 0.610 | -1.50 |
|  | HMGCLD | 14 | 22,930,291 | 0.061 | 0.002 | -30.55 | 0.019 | -3.24 |
|  | 3-MCC | 522 | 22,930,291 | 2.276 | 0.063 | -36.32 | 0.253 | -9.00 |
|  | MMA | 138 | 45,860,582 | 0.301 | 0.083 | -3.63 | 0.606 | 2.01 |
|  | PA | 125 | 22,930,291 | 0.545 | 0.009 | -60.44 | 0.687 | 1.26 |
|  | HCSD | 12 | 22,930,291 | 0.052 | 0.001 | -78.20 | 0.058 | 1.10 |
|  | BKT | 24 | 22,930,291 | 0.105 | 0.004 | -26.58 | 0.156 | 1.50 |
| Fatty Acid Oxidation Disorders | MCADD | 1,341 | 22,930,291 | 5.848 | 6.310 | 1.08 | 6.688 | 1.14 |
|  | LCHADD | 59 | 22,930,291 | 0.257 | 0.229 | -1.12 | 0.249 | -1.03 |
|  | VLCADD | 410 | 22,930,291 | 1.788 | 0.710 | -2.52 | 1.285 | -1.39 |
|  | TFPD | 14 | 22,930,291 | 0.061 | 0.230 | 3.77 | 0.274 | 4.48 |
|  | CDSP | 248 | 22,930,291 | 1.082 | 0.265 | -4.08 | 0.658 | -1.64 |
| Globinopathies | SCD+ | 11,593 | 22,930,291 | 50.558 | 1.762 | -28.70 | 2.486 | -20.34 |
| Endocrine Diseases | PCH | 14,388 | 22,930,291 | 62.747 | 2.849 | -22.02 | 5.210 | -12.04 |
|  | CAH | 1,591 | 22,930,291 | 6.938 | 24.511 | 3.53 | 20.794 | 3.00 |
| Lysosomal Diseases | GSD-II | 368 | 7,431,138 | 4.952 | 6.304 | 1.27 | 6.926 | 1.40 |
|  | MPS-I | 83 | 6,123,116 | 1.356 | 0.472 | -2.87 | 0.768 | -1.76 |
| Other | BIOT | 1,143 | 22,930,291 | 4.985 | 161.055 | 32.31 | 1.836 | -2.71 |
|  | CF | 4,969 | 22,719,414 | 21.871 | 32.771 | 1.50 | 21.075 | -1.04 |
|  | GALT | 472 | 22,930,291 | 2.058 | 1.023 | -2.01 | 1.440 | -1.43 |
|  | SMA | 219 | 3,185,560 | 6.875 | 0.093 | -73.75 | 0.631 | -10.89 |

### Genetic prevalence derived from gnomAD v4 is more concordant with NBS than gnomAD v2

We tested how the sample size of the genomic reference dataset used to estimate AF impacted our estimates of disease prevalence. While larger sample sizes should produce more accurate AF measures, additional samples may also capture singletons which could falsely inflate BP_g_.

We compared the concordance of BP_g_ from either gnomAD v2 (N = 141,456) or v4 (N = 807,162) versus BP_NBS_ and found a substantial increase in concordance in v4 compared to v2 (ρ_c_ = 0.61 vs 0.33, respectively) (**Figure S2**). The main reason appeared to be the poor estimate of SMA prevalence using gnomAD v2 (BP_NBS_ = 6.85 vs BP_g_ = 6.5×10^-6^), and concordance improves if SMA is removed (ρ_c_ = 0.64 and 0.75, V4 and V2 respectively). We speculate that this may result from reduced sequencing coverage of *SMN2* in the V2 dataset, quantified as ∼86% 10x dropout in V2 versus only ∼82% in V4 (**Figure S3**) (**Methods**). We further hypothesized underperformance of the remaining diseases in the V2 data may be due to differences in ancestry composition, population size, or sequencing methodology (**Figure S1**). Further comparison of gnomAD releases on estimation of AR genetic prevalence is provided in **Extended Dataset E1**. Given these results, we proceeded to refine the genetics-based prevalence estimates using the gnomAD v4 data exclusively.

### Filtering population-specific common alleles improves concordance

Prior reports suggest that ancestry-specific alleles with low clinical penetrance or severity can be over-represented in some populations due to reduced purifying selection.^55^ We hypothesized these were inflating the BP_g_ for some diseases identified as outliers (*e.g.*, BIOT) and that their removal would improve concordance with NBS. We compiled a list of 7 alleles with an allele frequency in any gnomAD ancestry group greater than 1% and annotated these for known pseudo-deficiency alleles (defined as mutations shown to have residual enzyme function and associated with mild or late-onset clinical disease) (Table S2).^23,56–59^ For example, in BIOT, we found an allele in the *BTD* gene (rs13078881) in nearly all ancestry groups (∼4% in European groups and ∼2% in others) that had been previously characterized as having low clinical penetrance in both homozygous and compound heterozygous individuals.^60^ We additionally found pseudo-deficiency alleles in both HBB (rs33930165; 1.5% in AFR), HBD (rs549964658; 2.6% in SAS), CYP21A2 (rs6471; 1.8% in AMR and 4.8% in ASJ), and CFTR (rs73715573; 1.5% in NFE). However, this strategy also removes known pathogenic alleles in HBB (rs334; 5% in AFR) and CFTR (rs73715573; >1% in nearly all groups), both of which are the major pathogenic variant for SCD and CF, respectively.

These findings suggested an additional revision to our analysis to remove population-specific common variants. Removal of these common alleles reduced diseases with overestimated BP_g_ (*eg*. BIOT from 32x overestimate to 13x underestimate), although at the expense of reducing concordance overall (ρ_c_ = 0.571 [95% CI 0.351-0.732]) (**Figure S4**). Given these results, we refined our genetics-based prevalence estimates by removing all alleles with >1% MAF for all future analyses.

### Correcting for ancestry-driven bias by modeling population demographics

Population ancestry stratification can significantly impact many genetic analyses^61,62^ and has been frequently incorporated into previous work on genetic prevalence.^23,24,26^ Next, we explored the impacts of ancestry on concordance between BP_g_ and BP_NBS_ in our data by stratifying the analysis by ancestry group (**Methods**). For this exploratory analysis, we switched to the California state (referred to as CA) NBS dataset because, unlike nationwide data, it includes parent self-reported ancestry for nearly all newborns.

Prior to ancestry stratification, we excluded seven diseases (HCU, HMGCLD, HCSD, BKT, TFPD, MPS-I, and SMA) with no reported cases in the CA data (**Extended Dataset E2**). Despite this, we noted the CA data was less concordant overall compared with our nationwide baseline (ρ_c_ = 0.526 [95% CI 0.218-0.739]) (**Figure S5A**). This reduced concordance could be due a significant shift in demographic composition of CA compared to gnomAD versus the nationwide dataset (Jensen–Shannon Divergence = 0.31 vs 0.09 bits, respectively; G-Test p < 10^-300^) (**Figure S1**).^63^

To test ancestry-specific concordance, we first mapped self-reported ethnicity from the CA NBS dataset to established gnomAD ancestry groups and calculated the CA aggregate and ancestry-specific BP_NBS_ (**Methods**). Since initial ancestry-stratified results were poor (**Extended Dataset E2**), we restricted our European-descent group to exclude Ashkenazim participants and replaced the Pan-Asian group with those of broad East Asian descent exclusively (**Methods**).

Despite these stricter ancestry-ethnicity matching rules, we observed concordance deteriorate across three of our four ancestry groups (ρ_c_ = 0.538 in White, 0.055 in Black, 0.342 in Admixed American, and 0.219 in Pan Asian) compared to CA in aggregate (ρ_c_ = 0.526) (**Figure S5**). Notably, there was high disease dropout in all ethnicity groups: 15/28 (54%) of diseases in White, 19/28 (68%) in Black, 10/28 (36%) in Admixed Americans, and 21/28 (75%) in Pan Asian (**Table S1**). The only demographic group with improved concordance, those categorized as European / white, was driven by collectively improved concordance (mean fold difference showed underestimate of 5x vs 7x in white vs aggregate, respectively).

We further hypothesized that matching the ancestry composition of gnomAD to that of the nationwide NBS data would improve the accuracy of genetic prevalence. To do this, we created a weighted sum of BP_g_ from gnomAD v4 ancestry groups based on U.S. census data (**Figure S1**). Applying this demographic weighting substantially improved concordance between BP_g_ and BP_NBS_ in our nationwide data (ρ_c_ = 0.75 [0.554-0.867 95% CI] versus 0.61 previously) with far fewer outliers observed (**Figure S6**). Specifically, 11/28 (39%) of diseases had a BP_g_ within 2x of BP_NBS_ and 24/28 (86%) were within 10x (**Table S1**). However, despite these improvements, the median genetic estimate remained ∼2.2-fold lower than that reported in NBS. One explanation for this result is that only a subset of all pathogenic variants has been included in ClinVar, leading to an underestimate of the true prevalence. Indeed, other groups have identified ClinVar P/LPs are unable to explain disease in most patients.^64^

### Inclusion of protein truncating, but not missense, variants predicted to be pathogenic further improves concordance

In an attempt to increase the coverage of pathogenic variants included in the model and further reduce the underestimate of prevalence compared to NBS, we tested whether including predicated pathogenic mutations improved genetic prevalence estimates. We focused on the inclusion of two classes of mutations that were not yet annotated as ClinVar P/LP: protein-truncating variants (PTVs) and missense variants predicted to be strongly protein damaging by *in silico* variant effect prediction (VEP) tools.

For the addition of missense mutations, we defined predicted pathogenic variants as those with an AlphaMissense (AM)^40^ score ≥ 0.575 or a REVEL^41^ score ≥ 0.7, the recommended cutoffs for both tools (**Methods**). Surprisingly, we found that inclusion of pathogenic predicated by AlphaMissene and REVEL decreased concordance between BP_g_ and BP_NBS_ (ρ_c_ = 0.451 [0.129-0.687]) (**Figure S7A**). Further, we found the median fold-difference is now 2x (**Extended Dataset E2**), potentially due to inclusion of non-pathogenic missense mutations inflating the estimate. For comparison, we also tested variants defined as pathogenic by AM and REVEL alone and found even worse concordance (ρ_c_ = 0.209 [-0.168-0.553]), suggesting inclusion of ClinVar P/LPs is critical to accurate estimation of BP_g_ (**Figure S7B**).

Next, we tested if addition of non-P/LP PTVs and found improved concordance between BP_g_ and BP_NBS_ (ρ_c_ = 0.79 [0.55-0.87 95% CI] versus 0.75 in the census-normalized analysis) (**Figure 1B**). The number of outliers was also further reduced with over half (16/28; 57%) of diseases with a BP_g_ within 2x of BPNBS and nearly all (25/28; 89%) within 10x (**Figure 1B-C**). The median fold difference narrowed to 1.4-fold lower than NBS, with only 3/28 (10%) of disease less than 10% of the NBS estimate (**Table 2**). The diseases that remained difficult to estimate (SMA, PCH, 3-MCC, SCD+) have at least one complicating attribute: (a) a known contribution by structural variants which we did not include,^65^ (b) a high degree of locus heterogeneity or mutations across multiple causal genes which were not considered in our model,^66^ (c) forms of the disease that remain silent or mild until adulthood without well elucidated predisposing genotype,^67^ or (d) diseases driven by alleles filtered out by our aggressive AF cutoff. Collectively, this series of straightforward filters and normalization approaches, resulted in a significant improvement in concordance from ρ_c_ = 0.61 in the baseline analysis to 0.79 in the final revised version.

## Discussion

Motivated by the importance of disease prevalence estimation for public health and pharmaceutical investment, we explored the viability of modeling AR disease prevalence from minimal genetic, epidemiological, and clinical data. We compared our estimates to NBS instead of epidemiology studies, which can suffer from ascertainment bias and non-quantitative diagnostic measures. NBS does not suffer from such limitations as most U.S. states report a participation rate of >99.9%^5^ and nearly all NBS assays employ quantitative measures of endophenotypes highly correlated with development of disease.^12–15^

Many genetics-first estimates reported in the literature are based on global allele frequencies, rather than stratifying by ancestry.^24,25,68–74^ Stratifying by ancestry improved some of the estimates, but worsened others. Normalizing the demographic composition of gnomAD to reflect the population structure of nationwide NBS data by utilizing U.S. census reports yielded estimates most concordant with NBS data, though this approach depends upon census data being available for the country or region of interest.

The impact of ancestry stratification on prevalence estimates is expected, given the regional specificity of genetic variation^75,76^ and of rare variants in particular.^77,78^ The ancestral composition of the data used to predict prevalence requires careful consideration, as does the demographic composition of the country being studied (for example, in the scenario where prevalence is being used to estimate the number of addressable patients in a given market). Even in small countries, rare variants can change in frequency across regions^79^ and genetic epidemiology studies have shown that disease-causing mutations can vary across continental groups, such as cystic fibrosis mutations across Europe,^80^ and even within countries, such as Friedreich’s Ataxia mutations across Spain.^81–83^

The disease mutation spectrum is also critical to estimating disease prevalence. Here, we have assumed the mutations we used capture the full disease mutation spectrum are fully penetrant, and that the genetic architecture of disease is limited to homozygous and compound heterozygous genotypes. However, very few diseases have a genetic architecture that is fully understood. Unidentified or misidentified mutations likely contribute to our systematic underestimate of disease prevalence; mutations may not yet be identified because genetic testing is still not part of the standard disease diagnostic pathway. For example, the American College of Medical Genetics (ACMG) recently recommended genetic testing be included in the diagnosis of PKU,^84^ a disease identified nearly a century ago, caused by >1,000 mutations, and for which genotype-phenotype relationships are still a subject of research.^85,86^ Due to the rareness of these diseases, generating cohorts of patients with available sequencing data is challenging, further confounding the complexities around interpreting genotype-phenotype relationships to identify improperly classified mutations; evaluating the potential pathogenicity of VUSs,^87,88^ for which a large number are reported for many rare diseases^89,90^; assessing mutation penetrance, which can be variable, even in rare disease^27,91,92^; and identifying potentially under-ascertained mutations, such as copy number or noncoding variants.^93,94^ We recommend, where possible, using disease-specific genotype-phenotype databases to further curate pathogenic mutations. Alternatively, VEP tools or frequency-based filters can be used to remove likely-benign variants from prevalence analyses.^95^

Finally, our prevalence estimates here make two additional assumptions that we have not probed further. The first assumption is that NBS data is the best way to measure birth prevalence of a disease. NBS data is not without limitations. Testing protocols across jurisdictions are not consistent, which hampers the reliability of nation-level data that aggregates results from multiple testing agencies, such as the U.S.-based dataset employed here. For example, differences in testing protocol or analyte diagnostic thresholds will yield variability in the sensitivity and specificity rates between centers. Further, although data captured under a single governing body or organization, such as state-level data, is more likely to utilize a consistent end-to-end methodology, there are additional sources of variance that should be recognized, such as changes in testing protocol over time or variation in public perceptions of self-reported ethnicity. The types of tests used to identify disease and testing efficacy itself also varies between diseases, complicating comparisons between birth prevalences.

Our second assumption is that we are studying disease burden in a randomly mating population. However, prevalence estimates should account for potential relatedness structure, especially when being applied to a part of the world with higher rates of consanguinity. Consanguinity can vary widely across cultures and countries, and even region to region within countries.^96,97^ Further, consanguinity is not rare; couples in consanguineous marriages are estimated to comprise >10% of the global population.^98^ Importantly, consanguinity can result in higher rates of disease^98^ and autosomal recessive disease is more common in consanguineous populations.^96–101^ For example, the autosomal recessive disease neuronal ceroid lipofuscinosis type 2 (CLN2) is rare and typically affects fewer than 1 in 100,000 individuals^102^; in Newfoundland, however, incidence is at 9 in 100,000. This elevated prevalence is driven in part by higher rates of consanguineous marriages in the province.^103^

While estimating disease birth prevalence from genetic data is affected by several factors and considerations, these estimates are not without value. The vast majority of rare diseases are not tested for on newborn screening panels.^104^ Therefore, even conservative genetic estimates can provide valuable information, including evaluating the accuracy of available disease epidemiology data generated via other methods. Further, there is a well-established bias in how research funding is distributed,^105,106^ and genetic estimates can inform potential disease epidemiology and burden in areas of the world not yet studied by classical epidemiology approaches. However, given the discrepancies we observe here between newborn screening data and estimates derived from allele frequencies, we still suggest caution when relying on genetics-based birth prevalence estimates. Considerations include the ancestral composition of the analyzed sample, the demographics and relatedness structure of the studied country, and understanding of the disease mutation spectrum. Estimates ignoring these caveats should be interpreted cautiously and in the context of disease epidemiology or newborn screening where available.

## Data Availability

All data produced in the present study are available upon reasonable request to the authors.

---

**Figure S1.**
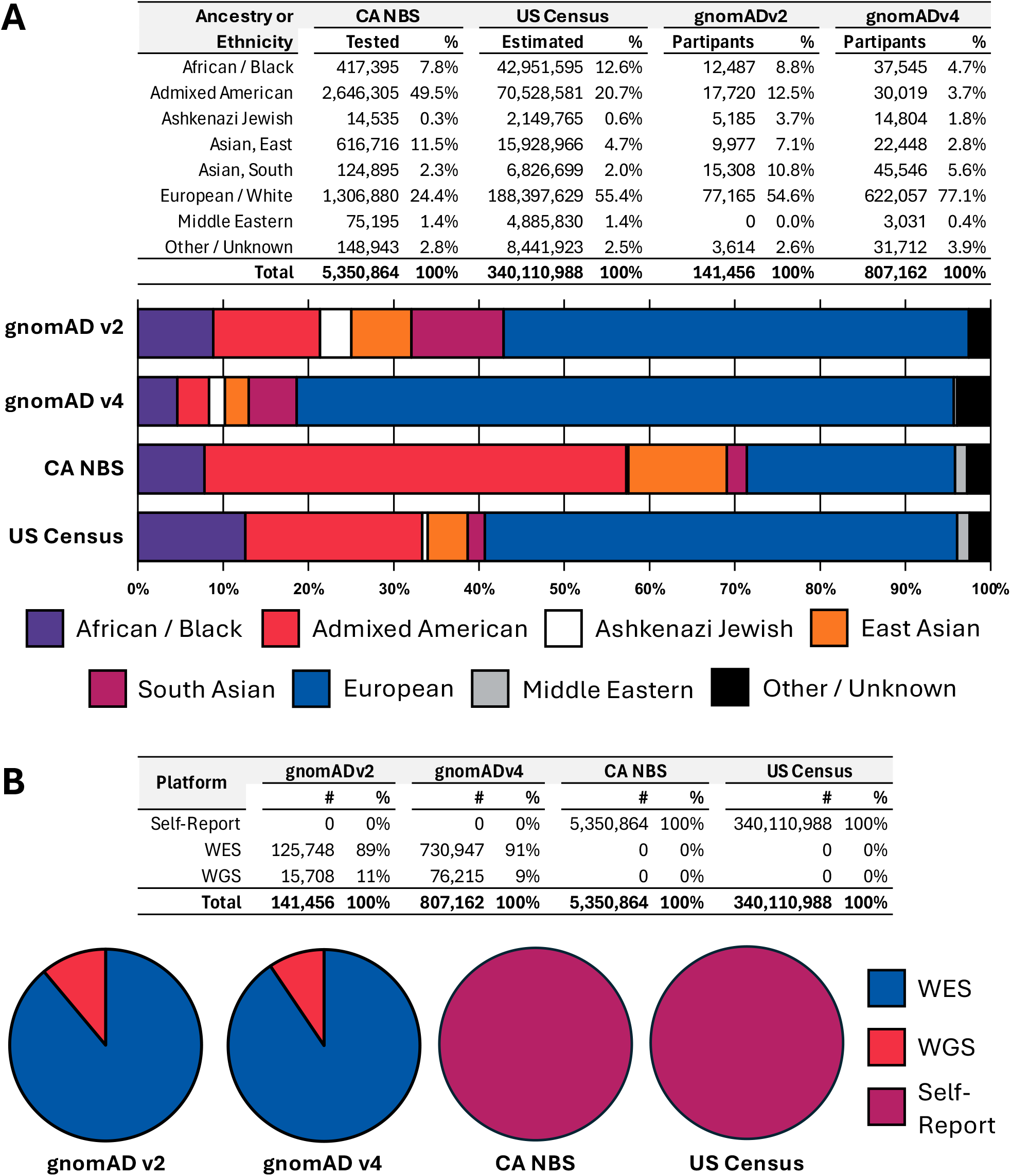
Comparison of dataset demographics. **[A]** Census data for 2024 was downloaded from the US Census Bureau. Additional demographic assumptions required to match self-identified ethnicity to gnomAD ancestral groups is provided in the Methods section. **[B]** Source of data for ethnicity data.

**Figure S2.**
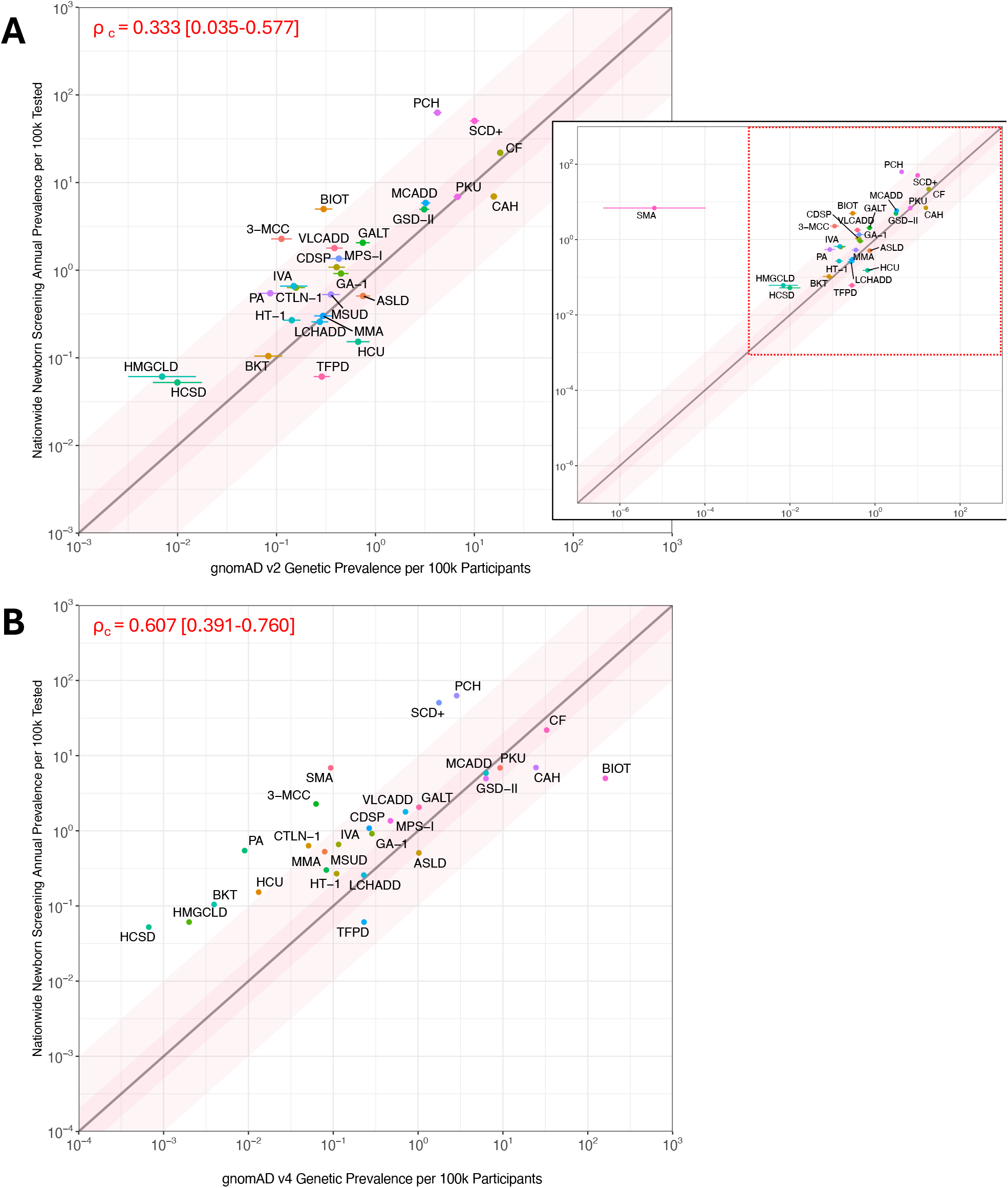
Concordance of autosomal recessive disease prevalence versus NBS is greater with genetic estimates derived from gnomAD v4 compared to gnomAD v2. **[A]** gnomADv2; inset includes wider range of prevalence values to highlight outliers. **[B]** gnomADv4

**Figure S3.**
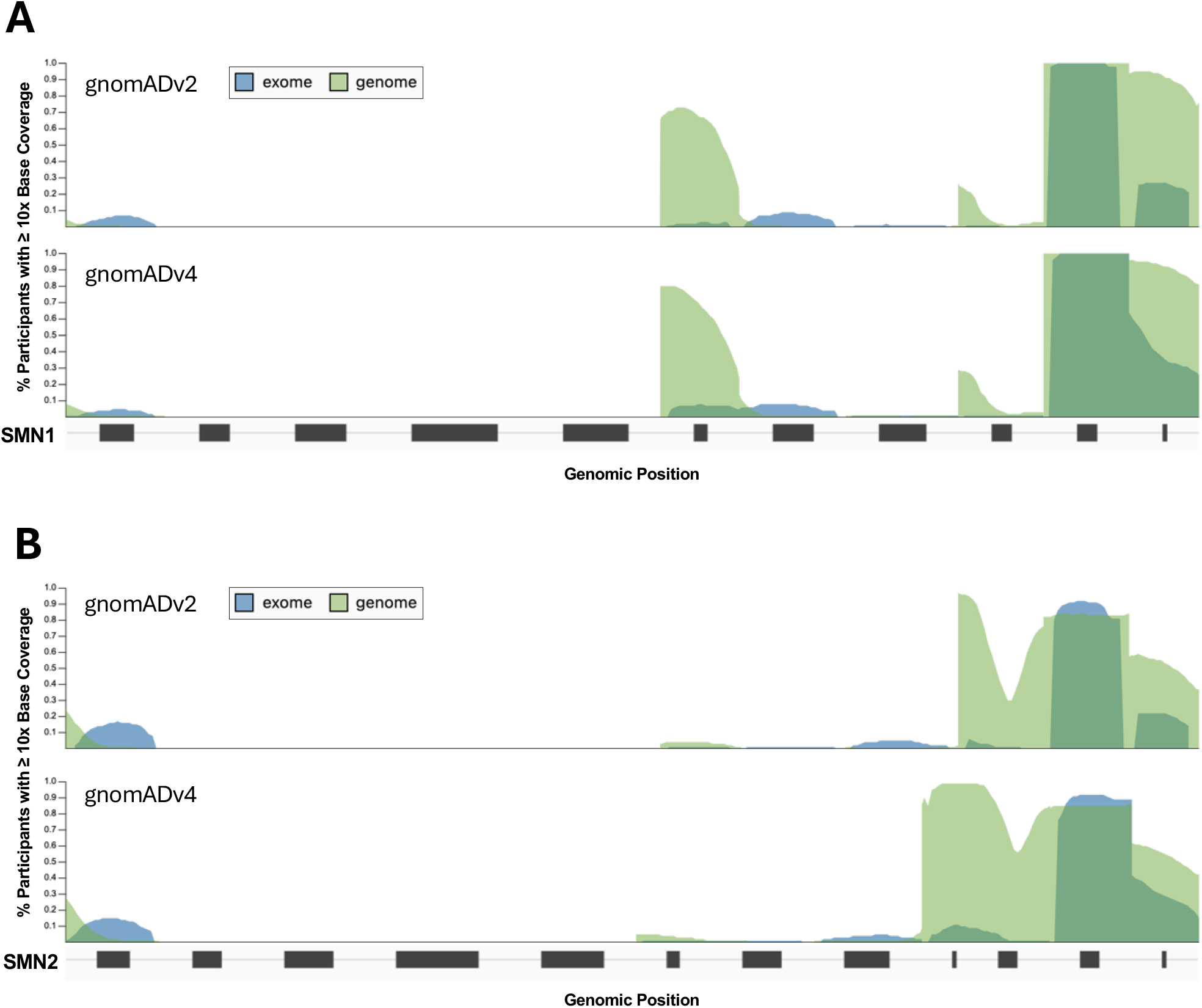
Sequencing coverage of SMN2 shows less target base dropout in gnomAD v4 versus v2. gnomAD sequencing coverage of **[A]** SMN1 **[B]** SMN2. Percent of samples covered ≥10x at a given base is shown on the Y-axis and base position in the gene body is shown on the X-axis, with exon shown schematically at the bottom. Samples with whole exome sequencing (WES) data is in blue and genome (WGS) in green.

**Figure S4.**
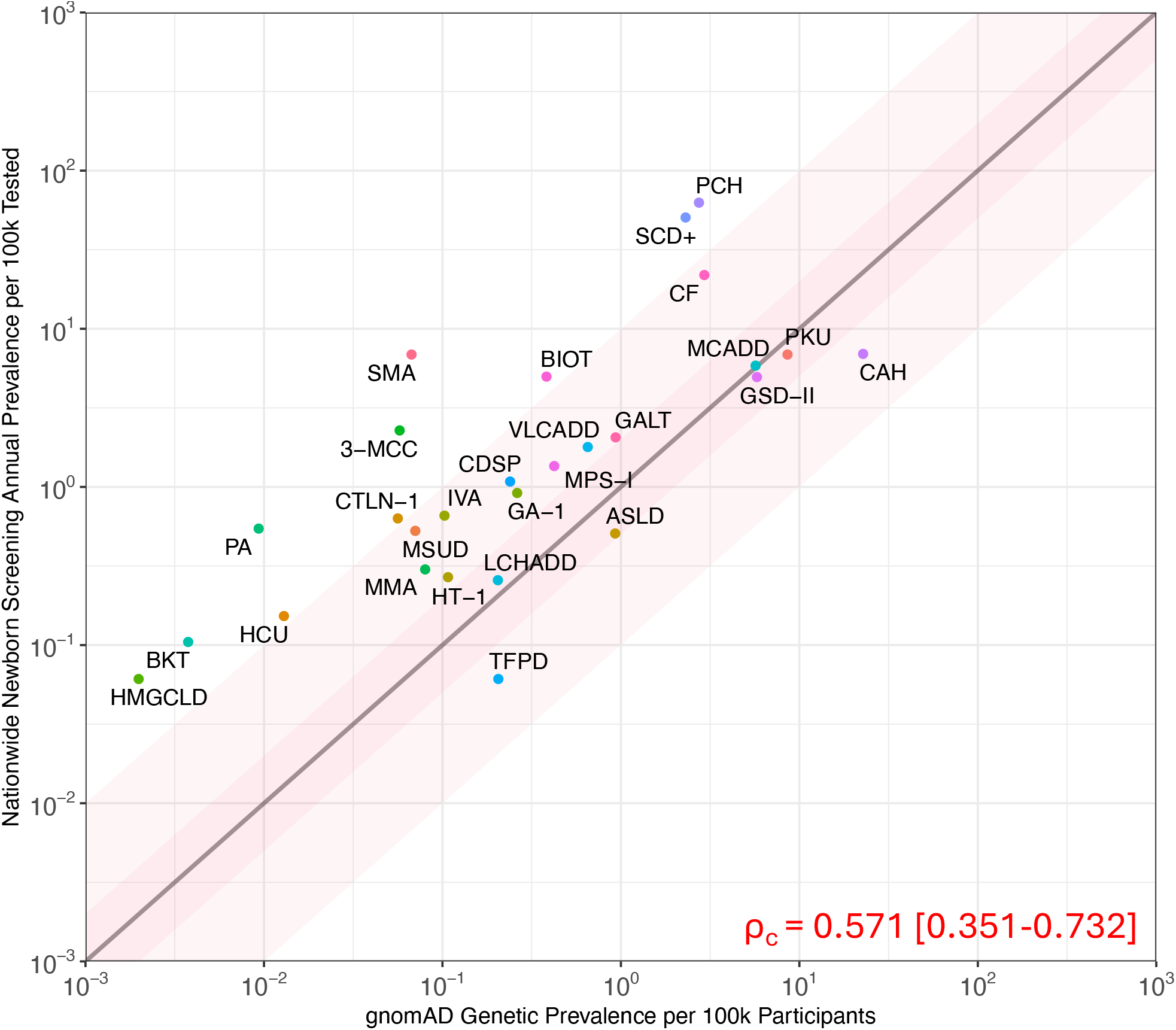
Removal of common pathogenic alleles in gnomAD removes overestimated outliers. Scatterplot of baseline genetic estimated prevalence (x-axis) versus the reported nationwide NBS prevalence (y-axis) for 28 AR diseases, both shown per 100k. The line of equality is shown in black with 2x and 10x ribbons in dark pink and light pink, respectively. Lin’s concordance correlation coefficient (ρ_c_) is annotated in red.

**Figure S5.**
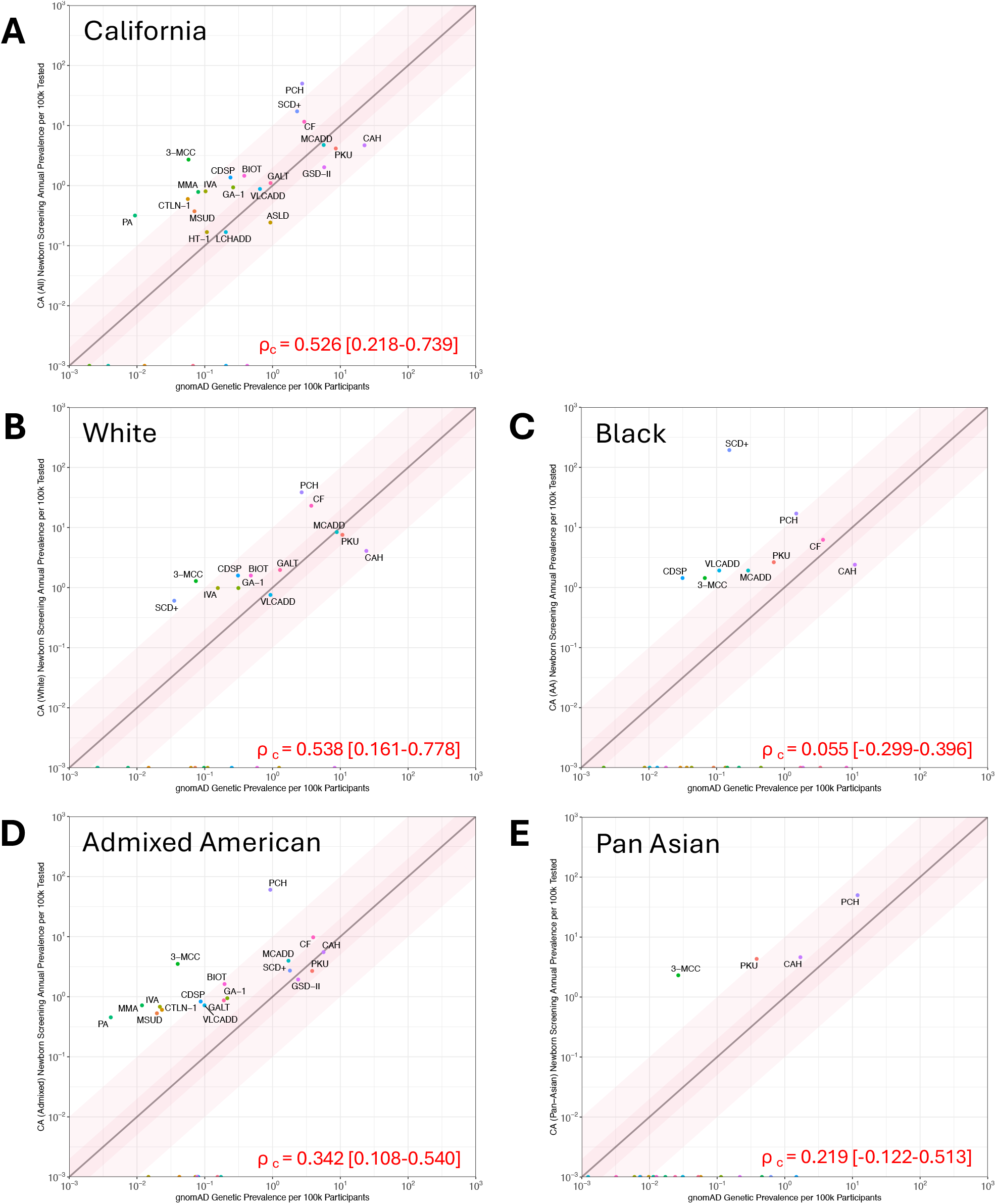
Stratification by ancestry does not yield improved concordance for AR disease prevalence. Scatterplot of genetic estimated prevalence versus the reported nationwide NBS prevalence for 28 AR diseases, both shown per 100k, for **[A]** the state of California (2009-2019) using all reported newborns and then this same data stratified by ancestry groups for **[B]** White, **[C]** Black, **[D]** Admixed American, and **[E]** Pan Asian. The line of equality is shown in black with 2x and 10x ribbons in dark pink and pink, respectively. Lin’s concordance correlation coefficient (ρ_c_) is annotated in red with 95% confidence intervals shown.

**Figure S6.**
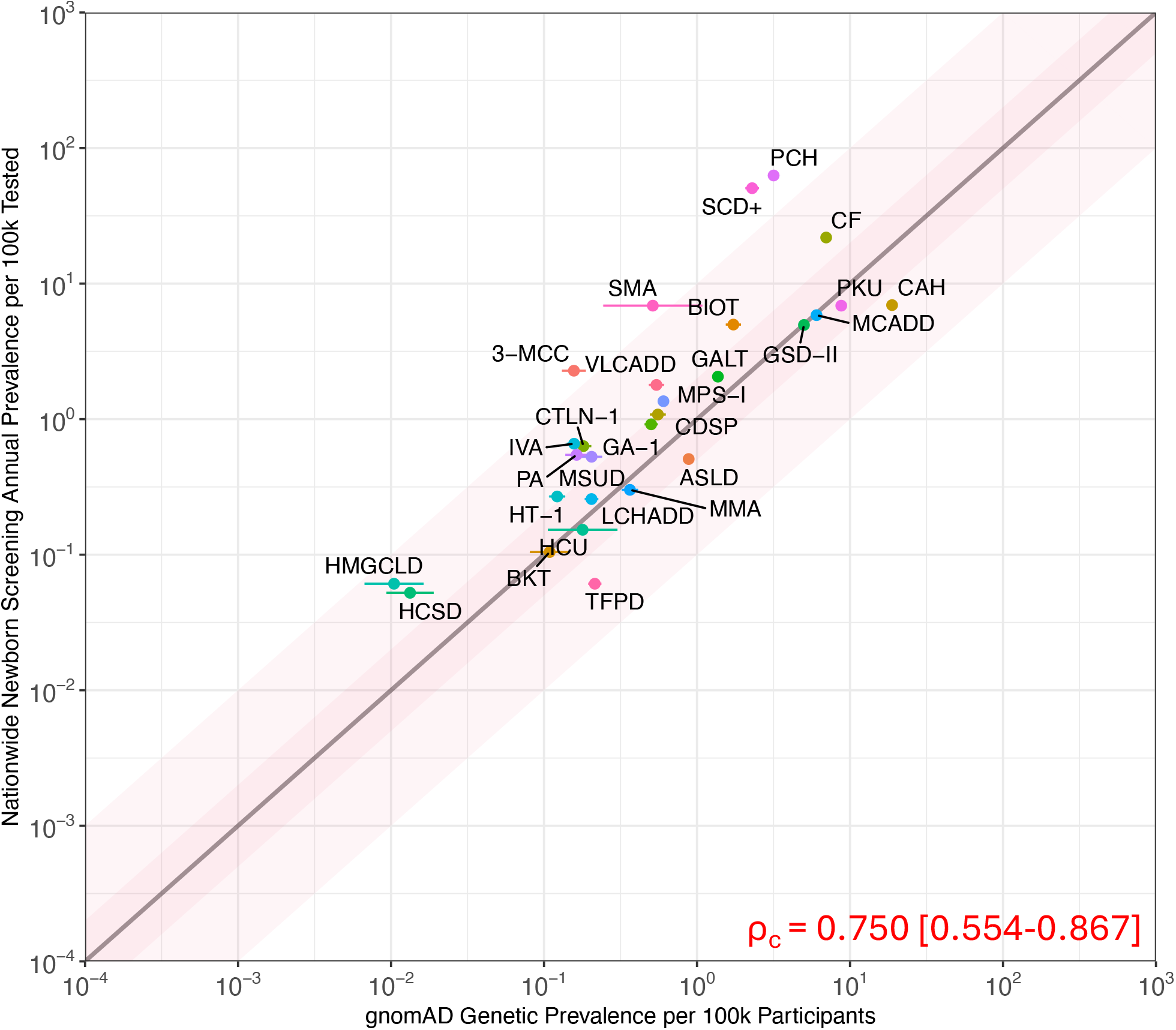
P_g_ vs P_NBS_ with census-adjusted ancestry normalization. Scatterplot of baseline genetic estimated prevalence (x-axis) versus the reported nationwide NBS prevalence (y-axis) for 28 AR diseases, both shown per 100k. The line of equality is shown in black with 2x and 10x ribbons in dark pink and light pink, respectively. Lin’s concordance correlation coefficient (ρ_c_) is annotated in red.

**Figure S7.**
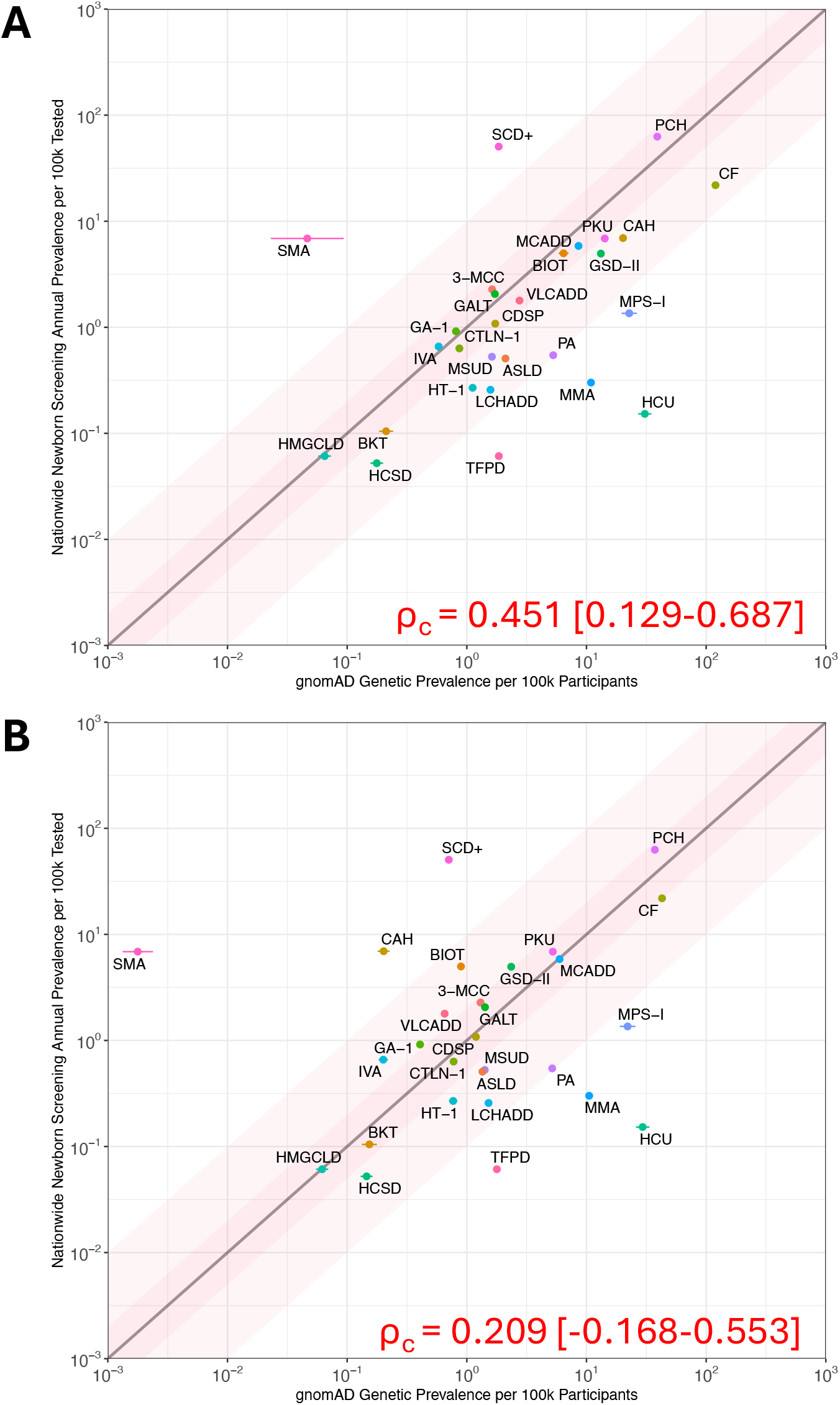
Incorporation of *in silico* predicted pathogenic variants reduces concordance between estimated genetic prevalence and NBS.

**Table S1.** Revisions to baseline model improve concordance and reduces outliers.

| Plot Labels |  | Lin's Rho<br>[95% C.I.] | Fold Difference to NBS |  |  |  |  |  |  |  |  |  |  |  |  |  |  |  |
| --- | --- | --- | --- | --- | --- | --- | --- | --- | --- | --- | --- | --- | --- | --- | --- | --- | --- | --- |
| Plot Description | Fig |  | Relative |  | Absolute |  | ± 2x |  | ± 10x |  | > 2x |  | > 10x |  | ≤ 2x |  | ≤ 10x |  |
|  |  |  | Median | Mean | Median | Mean | # | % | # | % | # | % | # | % | # | % | # | % |
| Nationwide Baseline (gnomADv2) | S1A | 0.333 [0.035-0.577] | -2.36 | -37,828.63 | 2.97 | 37,829.62 | 10 | 36% | 24 | 86% | 13 | 46% | 0 | 0% | 25 | 89% | 28 | 100% |
| Nationwide Baseline (gnomADv4) | 1A | 0.607 [0.391-0.760] | -3.42 | -13.14 | 3.93 | 16.48 | 5 | 18% | 17 | 61% | 9 | 32% | 1 | 4% | 24 | 86% | 27 | 96% |
| Nationwide + AF Filter | S4 | 0.571 [0.351-0.732] | -5.46 | -16.56 | 5.46 | 17.34 | 5 | 18% | 17 | 61% | 7 | 25% | 0 | 0% | 26 | 93% | 28 | 100% |
| CA + AF Filter | S5A | 0.491 [0.170-0.717] | -2.23 | -3.52 | 2.65 | 4.58 | 11 | 39% | 24 | 86% | 13 | 46% | 0 | 0% | 26 | 93% | 28 | 100% |
| CA + AF Filter [White - original] | NA | 0.461 [0.074-0.728] | -3.81 | -6.93 | 3.94 | 8.45 | 5 | 18% | 17 | 61% | 9 | 32% | 0 | 0% | 17 | 61% | 21 | 75% |
| CA + AF Filter [Black - original] | NA | 0.581 [0.201-0.809] | -3.58 | -5.45 | 5.22 | 6.72 | 4 | 14% | 10 | 36% | 5 | 18% | 0 | 0% | 12 | 43% | 13 | 46% |
| CA + AF Filter [Admixed - original] | NA | 0.334 [0.077-0.549] | -11.38 | -151.53 | 11.38 | 152.55 | 1 | 4% | 4 | 14% | 2 | 7% | 0 | 0% | 8 | 29% | 9 | 32% |
| CA + AF Filter [Asian - original] | NA | 0.209 [-0.109-0.489] | -7.77 | -24.71 | 7.77 | 25.12 | 4 | 14% | 11 | 39% | 4 | 14% | 0 | 0% | 18 | 64% | 18 | 64% |
| CA + AF Filter [White - revised] | S5B | 0.461 [0.074-0.728] | -8.07 | -1,506.28 | 8.07 | 1,506.60 | 2 | 7% | 4 | 14% | 2 | 7% | 0 | 0% | 7 | 25% | 7 | 25% |
| CA + AF Filter [Black - revised] | S5C | 0.581 [0.201-0.809] | -3.32 | -4.99 | 5.14 | 6.47 | 4 | 14% | 10 | 36% | 5 | 18% | 0 | 0% | 12 | 43% | 13 | 46% |
| CA + AF Filter [Admixed - revised] | S5D | 0.334 [0.077-0.549] | -11.38 | -151.53 | 11.38 | 152.55 | 1 | 4% | 4 | 14% | 2 | 7% | 0 | 0% | 8 | 29% | 9 | 32% |
| CA + AF Filter [Asian - revised] | S5E | 0.209 [-0.109-0.489] | -7.77 | -24.71 | 7.77 | 25.12 | 4 | 14% | 11 | 39% | 4 | 14% | 0 | 0% | 18 | 64% | 18 | 64% |
| Nationwide + AF Filter + Normalized | S4 | 0.750 [0.554-0.867] | -8.07 | -1,506.28 | 8.07 | 1,506.60 | 2 | 7% | 4 | 14% | 2 | 7% | 0 | 0% | 7 | 25% | 7 | 25% |
| Nationwide + AF Filter + Normalized + VUS | S7A | 0.451 [0.129-0.687] | 2.04 | 5.60 | 2.79 | 18.59 | 11 | 39% | 22 | 79% | 26 | 93% | 4 | 14% | 13 | 46% | 24 | 86% |
| Nationwide + AF Filter + Normalized + VUS - P/LPs | S7B | 0.209 [-0.168-0.553] | 1.06 | -131.68 | 2.71 | 153.65 | 10 | 36% | 21 | 75% | 20 | 71% | 4 | 14% | 18 | 64% | 24 | 86% |
| Nationwide + AF Filter + Normalized + VUS + PTVs | 1B | 0.792 [0.554-0.867] | -1.41 | -1.85 | 1.78 | 3.55 | 16 | 57% | 25 | 89% | 20 | 71% | 0 | 0% | 24 | 86% | 28 | 100% |

‘Relative’ means the original fold difference (FD) values that are relative (+/-) to NBS. ‘Absolute’ means the absolute value of all FDs are taken.

**Table S2.**
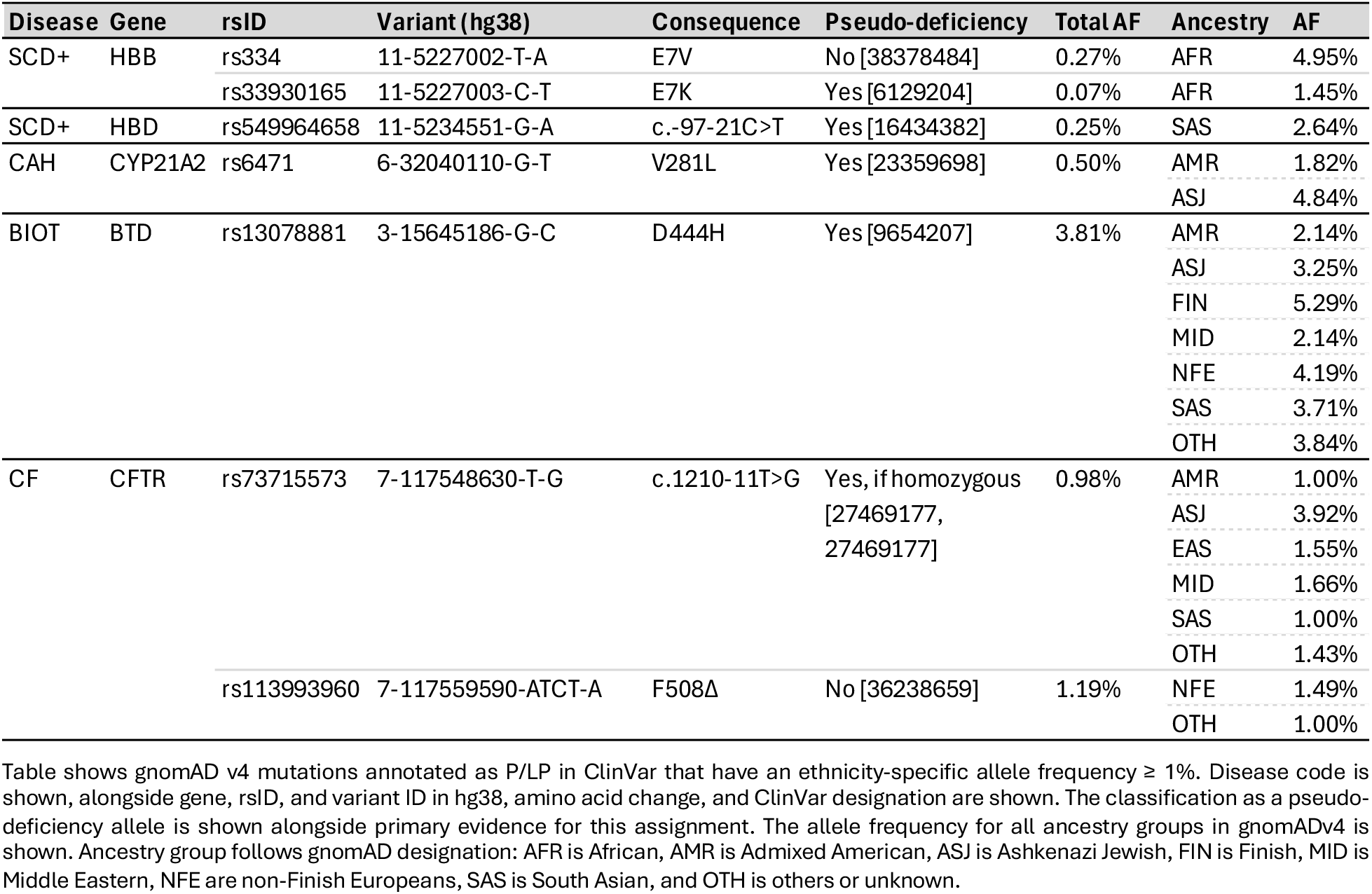
Examples of alleles with high ancestry-specific frequency and clinical effect on protein function.

