## Extended Datasets for "Benchmarking autosomal recessive disease prevalence estimation from allele frequencies against newborn screening data"

**Extended Dataset E1: Effect of source genetic database by comparing gnomAD v2 to v4**

We tested how the sample size of the reference dataset used to estimate allele frequencies impacted our estimates of disease prevalence. The Genome Aggregation Database (gnomAD) has grown from version 2.1.1 (N=141,456 individuals: 125,748 exomes, 15,708 genomes) to version 4.1 (N=807,162 individuals: 730,947 exomes, 76,215 genomes) — a 5.7-fold increase across more than nine ancestral groups.[3,5] We examined how this expansion alters BP predictions with the goal of comparing to real-world incidence data.

See **Methods** for additional description, but in brief: Under Hardy-Weinberg equilibrium[4], prevalence per 100,000 is calculated as BP = 1 − product over i of (1 − f_i)^2 × 10^5. Ancestries are weighted by their proportion in the population. Uncertainty is estimated using the delta method. Gene-level tests use a two-sided Wald test on the logit scale with Benjamini-Hochberg FDR correction.[1] The global v2 to v4 fit uses an errors-in-variables (York) model on the logit scale: logit(theta_4) = alpha + beta × theta_2, where theta = logit(prevalence/10^5). Linear fits are also reported. Fit criteria minimize chi-squared error, accounting for correlated errors.

We compared gene-level prevalence estimates between gnomAD v2.1.1 and v4.1 using weighted ancestry composition, per-ancestry 1% allele frequency filter, ClinVar Pathogenic or Likely Pathogenic variants, and disease genes as defined by Choi et al.[2] Pearson correlation was 0.914, Spearman correlation was 0.921. The York (logit) fit was alpha = -0.221, beta = 0.990. The distribution of log2(v4/v2) fold changes per ancestry showed that most genes had similar prevalence estimates between versions, but some genes had notable differences. The median fold change per ancestry varied.

Of 286 genes, 26 (9.1%) were significantly different between v2 and v4 after FDR correction. 20 genes had lower prevalence in v4 (the most common pattern), while 6 genes had higher prevalence in v4 (PREPL, ADSS1, TPP1, CHRNG, PYGM, GAA). PREPL showed the most significant increase (8.2-fold, p adjusted = 1.4e-16). Across ancestry groups, the pooled median v4/v2 fold change was 0.89. Most ancestry groups showed similar trends, but some genes and populations had larger differences.

Of 286 genes, 26 (9.1%) were significantly different between v2 and v4. 20 genes were lower in v4, 6 genes were higher. PREPL had the most significant increase. Pearson correlation was 0.914, Spearman correlation was 0.921. The fitted v2 to v4 relation per 100,000 was $prev\_4 \approx -0.000 + 0.676 \times prev\_2$. The logit fit was $theta\_4 \approx -0.221 + 0.990 \times theta\_2$, implying a multiplicative change on the odds scale.

Coverage is not uniform across the genome; many genes and regions remain low-coverage in both v2 and v4. Sensitivity at low coverage means small allele frequency changes can produce large shifts in gene-level prevalence, so estimates can be volatile. The central tendency is stable, with exceptions: on average, v4 tracks v2, but numerous genes show meaningful differences after FDR control. Bigger sample size helps but isn't sufficient; durable improvements require better coverage, ancestry balance, and variant curation/QC alongside transparent uncertainty reporting.

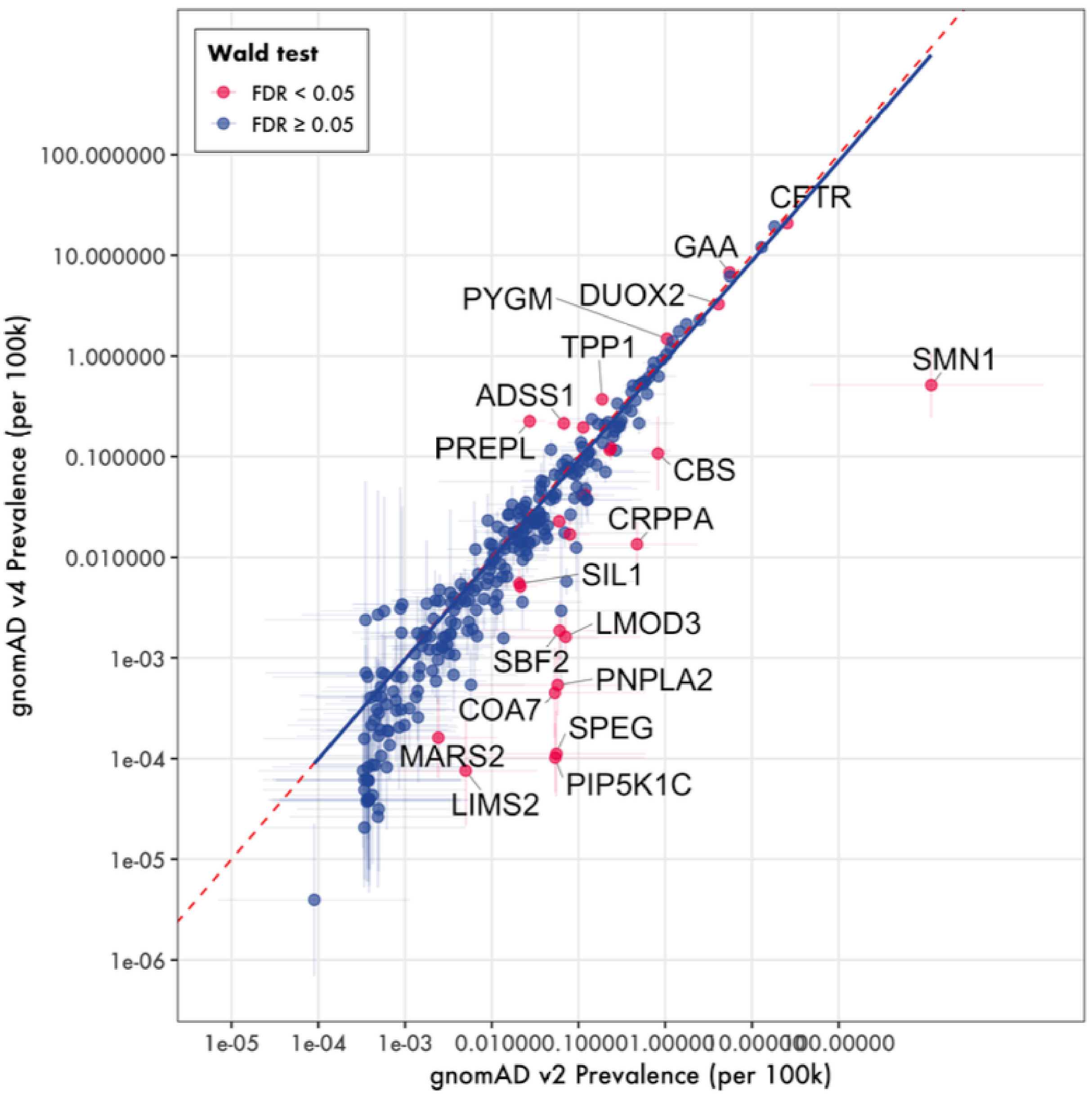


**Figure E1: Gene-Level Prevalence Comparison**

Gene-level prevalence v2 vs v4 (N=286 genes, 26 significant at FDR ¡ 0.05). Error bars show 95% CIs (logit). Pearson r = 0.914, Spearman ρ= 0.921. York (logit) fit: α = -0.221, β = 0.990.

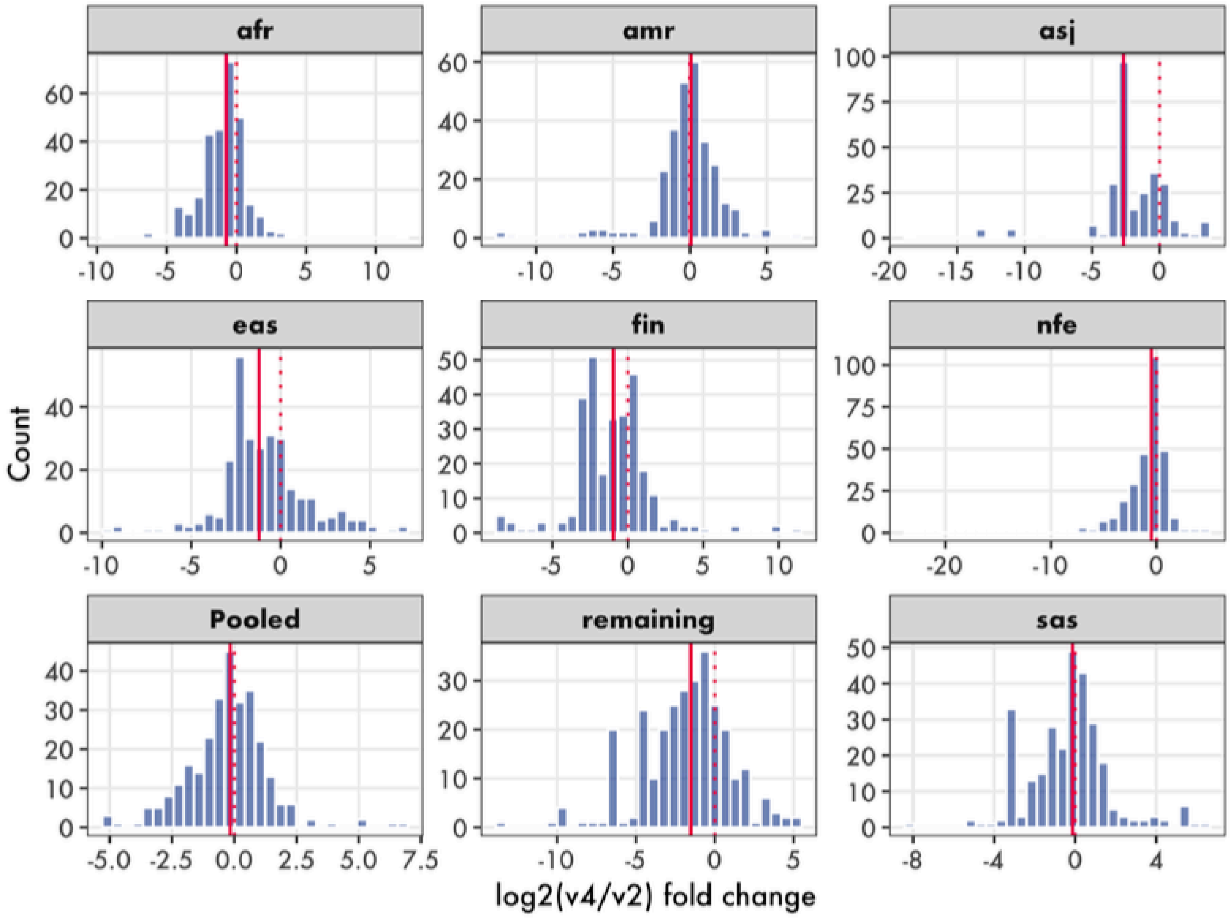


**Figure E2: Distribution of prevalence estimate differences by ancestry**

Distribution of log2(v4/v2) fold changes per ancestry. Dotted line: no change (0); solid line: median per ancestry.

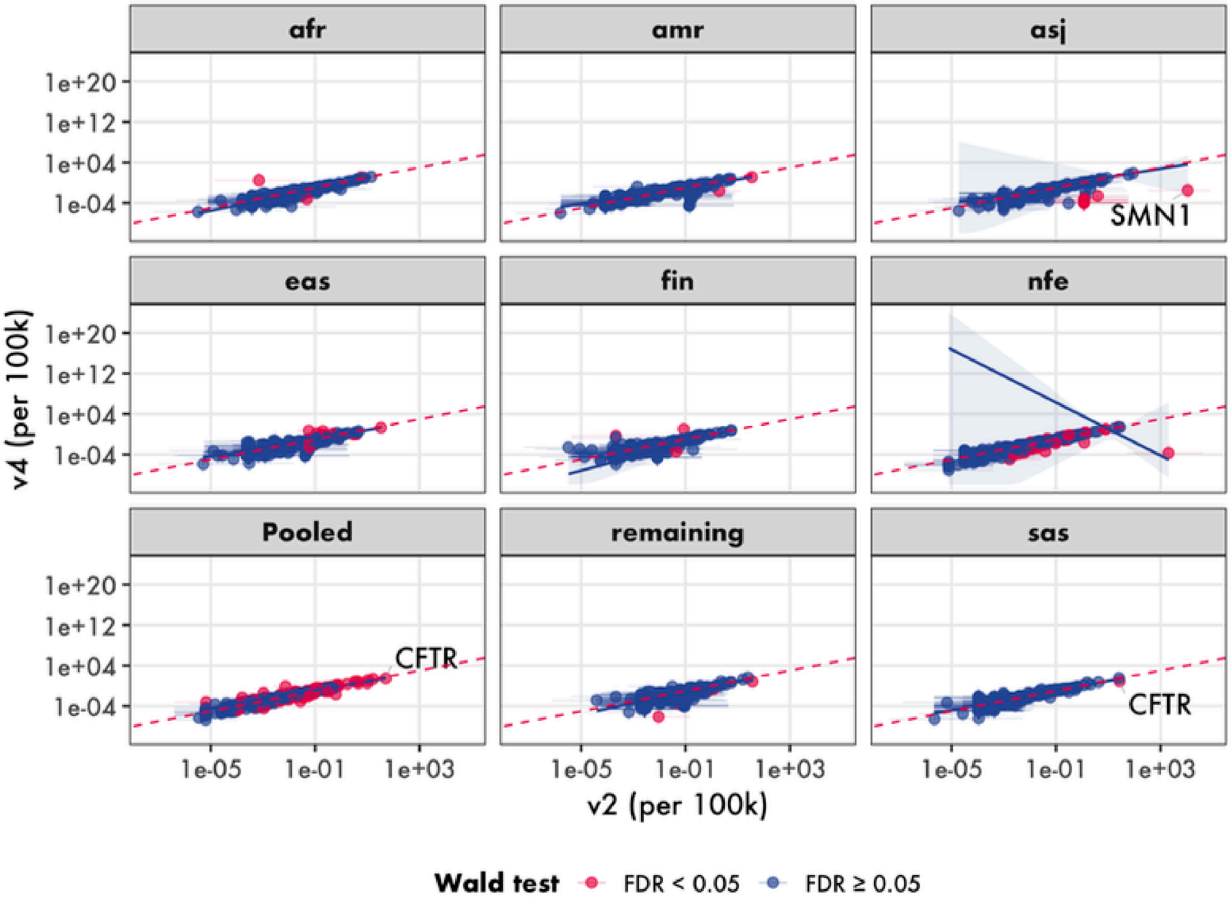


**Figure 3: Prevalence estimate differences by ancestry at gene-level**

Gene-level prevalence v4 vs v2 per ancestry (log-log scales). Top 3 significant genes per ancestry labeled (FDR <0.05). Error bars show 95% CIs.

**Extended Dataset E2 [Part 1 of 3]**

| | | | Nationwide | | | | | | | | | | | | | | | | |
|---|---|---|---|---|---|---|---|---|---|---|---|---|---|---|---|---|---|---|---|
| | | | | | | Genetic Estimate | | | | | | | | | | | | | |
| | Therapeutic Area | Disease Code | NBS | | | gnomAD v2 P/LPs N/A Aggregate | | gnomAD v4 P/LPs N/A Aggregate | | gnomAD v4 P/LPs ≤ 1% Aggregate | | gnomAD v4 P/LPs ≤ 1% Census-adjusted | | gnomAD v4 P/LPs + Pred_Mis ≤ 1% Census-adjusted | | gnomAD v4 PTVs + Pred_Mis ≤ 1% Census-adjusted | | gnomAD v4 P/LPs + PTVs ≤ 1% Census-adjusted | |
| | | | **Cases** | **Births** | **:100k** | **:100k** | **FD** | **:100k** | **FD** | **:100k** | **FD** | **:100k** | **FD** | **:100k** | **FD** | **:100k** | **FD** | **:100k** | **FD** |
| 1 | Amino Acid Disorders | PKU | 1,576 | 22,930,291 | 6.873 | 6.743 | -1.02 | 9.256 | 1.35 | 8.581 | 1.25 | 8.770 | 1.28 | 14.214 | 2.07 | 5.232 | -1.31 | 12.168 | 1.77 |
| 2 | | MSUD | 121 | 22,930,291 | 0.528 | 0.355 | -1.49 | 0.079 | -6.66 | 0.070 | -7.49 | 0.205 | -2.58 | 1.620 | 3.07 | 1.413 | 2.68 | 0.349 | -1.51 |
| 3 | | HCU | 35 | 22,930,291 | 0.153 | 0.667 | 4.37 | 0.013 | -11.56 | 0.013 | -11.80 | 0.179 | 1.17 | 30.722 | 201.27 | 29.459 | 193.00 | 0.635 | 4.16 |
| 4 | | CTLN-1 | 145 | 22,930,291 | 0.632 | 0.157 | -4.03 | 0.051 | -12.36 | 0.056 | -11.25 | 0.181 | -3.50 | 0.867 | 1.37 | 0.775 | 1.23 | 0.311 | -2.03 |
| 5 | | ASLD | 116 | 22,839,659 | 0.508 | 0.750 | 1.48 | 1.020 | 2.01 | 0.929 | 1.83 | 0.882 | 1.74 | 2.103 | 4.14 | 1.350 | 2.66 | 0.996 | 1.96 |
| 6 | | HT-1 | 61 | 22,695,225 | 0.269 | 0.143 | -1.88 | 0.109 | -2.46 | 0.107 | -2.50 | 0.122 | -2.20 | 1.119 | 4.16 | 0.769 | 2.86 | 0.150 | -1.79 |
| 7 | Organic Acid Disorders | IVA | 151 | 22,930,291 | 0.659 | 0.149 | -4.41 | 0.116 | -5.70 | 0.103 | -6.41 | 0.157 | -4.19 | 0.581 | -1.13 | 0.200 | -3.29 | 0.301 | -2.19 |
| 8 | | GA-1 | 210 | 22,930,291 | 0.916 | 0.448 | -2.04 | 0.285 | -3.21 | 0.262 | -3.49 | 0.502 | -1.83 | 0.811 | -1.13 | 0.407 | -2.25 | 0.610 | -1.50 |
| 9 | | HMGCLD | 14 | 22,930,291 | 0.061 | 0.007 | -8.76 | 0.002 | -30.55 | 0.002 | -30.76 | 0.010 | -5.84 | 0.065 | 1.06 | 0.062 | 1.01 | 0.019 | -3.24 |
| 10 | | 3-MCC | 522 | 22,930,291 | 2.276 | 0.112 | -20.30 | 0.063 | -36.32 | 0.058 | -39.54 | 0.157 | -14.52 | 1.626 | -1.40 | 1.302 | -1.75 | 0.253 | -9.00 |
| 11 | | MMA | 138 | 45,860,582 | 0.301 | 0.296 | -1.02 | 0.083 | -3.63 | 0.080 | -3.76 | 0.364 | 1.21 | 10.908 | 36.25 | 10.528 | 34.99 | 0.606 | 2.01 |
| 12 | | PA | 125 | 22,930,291 | 0.545 | 0.086 | -6.31 | 0.009 | -60.44 | 0.009 | -58.38 | 0.163 | -3.34 | 5.271 | 9.67 | 5.172 | 9.49 | 0.687 | 1.26 |
| 13 | | HCSD | 12 | 22,930,291 | 0.052 | 0.010 | -5.26 | 0.001 | -78.20 | 0.001 | -88.65 | 0.013 | -3.93 | 0.177 | 3.38 | 0.145 | 2.78 | 0.058 | 1.10 |
| 14 | | BKT | 24 | 22,930,291 | 0.105 | 0.083 | -1.27 | 0.004 | -26.58 | 0.004 | -27.83 | 0.109 | 1.04 | 0.212 | 2.02 | 0.153 | 1.47 | 0.156 | 1.50 |
| 15 | Fatty Acid Oxidation Disorders | MCADD | 1,341 | 22,930,291 | 5.848 | 3.221 | -1.82 | 6.310 | 1.08 | 5.684 | -1.03 | 6.043 | 1.03 | 8.577 | 1.47 | 5.958 | 1.02 | 6.688 | 1.14 |
| 16 | | LCHADD | 59 | 22,930,291 | 0.257 | 0.274 | 1.06 | 0.229 | -1.12 | 0.204 | -1.26 | 0.204 | -1.26 | 1.576 | 6.12 | 1.519 | 5.90 | 0.249 | -1.03 |
| 17 | | VLCADD | 410 | 22,930,291 | 1.788 | 0.386 | -4.64 | 0.710 | -2.52 | 0.652 | -2.74 | 0.543 | -3.29 | 2.757 | 1.54 | 0.653 | -2.74 | 1.285 | -1.39 |
| 18 | | TFPD | 14 | 22,930,291 | 0.061 | 0.287 | 4.70 | 0.230 | 3.77 | 0.206 | 3.37 | 0.215 | 3.52 | 1.851 | 30.31 | 1.783 | 29.20 | 0.274 | 4.48 |
| 19 | | CDSP | 248 | 22,930,291 | 1.082 | 0.405 | -2.67 | 0.265 | -4.08 | 0.239 | -4.52 | 0.555 | -1.95 | 1.728 | 1.60 | 1.191 | 1.10 | 0.658 | -1.64 |
| 20 | Globinopathies | SCD+ | 11,593 | 22,930,291 | 50.558 | 10.039 | -5.04 | 1.762 | -28.70 | 2.304 | -21.94 | 2.293 | -22.05 | 1.846 | -27.39 | 0.707 | -71.47 | 2.486 | -20.34 |
| 21 | Endocrine Diseases | PCH | 14,388 | 22,930,291 | 62.747 | 4.230 | -14.84 | 2.849 | -22.02 | 2.738 | -22.92 | 3.168 | -19.81 | 39.055 | -1.61 | 37.257 | -1.68 | 5.210 | -12.04 |
| 22 | | CAH | 1,591 | 22,930,291 | 6.938 | 15.722 | 2.27 | 24.511 | 3.53 | 22.760 | 3.28 | 18.839 | 2.72 | 20.193 | 2.91 | 0.202 | -34.38 | 20.794 | 3.00 |
| 23 | Lysosomal Diseases | GSD-II | 368 | 7,431,138 | 4.952 | 3.112 | -1.59 | 6.304 | 1.27 | 5.785 | 1.17 | 5.003 | 1.01 | 13.187 | 2.66 | 2.348 | -2.11 | 6.926 | 1.40 |
| 24 | | MPS-I | 83 | 6,123,116 | 1.356 | 0.426 | -3.18 | 0.472 | -2.87 | 0.423 | -3.20 | 0.603 | -2.25 | 22.754 | 16.79 | 22.056 | 16.27 | 0.768 | -1.76 |
| 25 | Other | BIOT | 1,143 | 22,930,291 | 4.985 | 0.299 | -16.67 | 161.055 | 32.31 | 0.383 | -13.02 | 1.729 | -2.88 | 6.431 | 1.29 | 0.893 | -5.58 | 1.836 | -2.71 |
| 26 | | CF | 4,969 | 22,719,414 | 21.871 | 18.248 | -1.20 | 32.771 | 1.50 | 2.934 | -7.45 | 6.986 | -3.13 | 119.707 | 5.47 | 42.787 | 1.96 | 21.075 | -1.04 |
| 27 | | GALT | 472 | 22,930,291 | 2.058 | 0.745 | -2.76 | 1.023 | -2.01 | 0.934 | -2.20 | 1.369 | -1.50 | 1.717 | -1.20 | 1.418 | -1.45 | 1.440 | -1.43 |
| 28 | | SMA | 219 | 3,185,560 | 6.875 | 0.000 | -1,059,103 | 0.093 | -73.75 | 0.067 | -102.43 | 0.514 | -13.37 | 0.046 | -148.08 | 0.002 | -3,866.60 | 0.631 | -10.89 |

**Extended Dataset E2 [Part 2 of 3]**

| | Therapeutic Area | Disease Code | California State NBS Data | | | | | | | | | | | | | | |
|---|---|---|---|---|---|---|---|---|---|---|---|---|---|---|---|---|---|
| | | | CA NBS | | | | | | | | | | | | | | |
| | | | Aggregate (All) | | | White / European | | | Black / African | | | Admixed American | | | Pan-Asian | | |
| | | | Cases | Births | :100k | Cases | Births | :100k | Cases | Births | :100k | Cases | Births | :100k | Cases | Births | :100k |
| 1 | Amino Acid Disorders | PKU | 222 | 5,350,864 | 4.149 | 100 | 1,321,415 | 7.568 | 11 | 417,395 | 2.635 | 71 | 2,646,305 | 2.683 | 23 | 741,611 | 3.101 |
| 2 | | MSUD | 20 | 5,350,864 | 0.374 | 0 | 1,321,415 | 0.000 | 0 | 417,395 | 0.000 | 14 | 2,646,305 | 0.529 | 0 | 741,611 | 0.000 |
| 3 | | HCU | 0 | 5,350,864 | 0.000 | 0 | 1,321,415 | 0.000 | 0 | 417,395 | 0.000 | 0 | 2,646,305 | 0.000 | 0 | 741,611 | 0.000 |
| 4 | | CTLN-1 | 32 | 5,350,864 | 0.598 | 0 | 1,321,415 | 0.000 | 0 | 417,395 | 0.000 | 16 | 2,646,305 | 0.605 | 0 | 741,611 | 0.000 |
| 5 | | ASLD | 13 | 5,350,864 | 0.243 | 0 | 1,321,415 | 0.000 | 0 | 417,395 | 0.000 | 0 | 2,646,305 | 0.000 | 0 | 741,611 | 0.000 |
| 6 | | HT-1 | 9 | 5,350,864 | 0.168 | 0 | 1,321,415 | 0.000 | 0 | 417,395 | 0.000 | 0 | 2,646,305 | 0.000 | 0 | 741,611 | 0.000 |
| 7 | Organic Acid Disorders | IVA | 43 | 5,350,864 | 0.804 | 13 | 1,321,415 | 0.984 | 0 | 417,395 | 0.000 | 18 | 2,646,305 | 0.680 | 0 | 741,611 | 0.000 |
| 8 | | GA-1 | 50 | 5,350,864 | 0.934 | 13 | 1,321,415 | 0.984 | 0 | 417,395 | 0.000 | 25 | 2,646,305 | 0.945 | 0 | 741,611 | 0.000 |
| 9 | | HMGCLD | 0 | 5,350,864 | 0.000 | 0 | 1,321,415 | 0.000 | 0 | 417,395 | 0.000 | 0 | 2,646,305 | 0.000 | 0 | 741,611 | 0.000 |
| 10 | | 3-MCC | 145 | 5,350,864 | 2.710 | 17 | 1,321,415 | 1.286 | 6 | 417,395 | 1.437 | 93 | 2,646,305 | 3.514 | 8 | 741,611 | 1.079 |
| 11 | | MMA | 42 | 5,350,864 | 0.785 | 0 | 1,321,415 | 0.000 | 0 | 417,395 | 0.000 | 19 | 2,646,305 | 0.718 | 0 | 741,611 | 0.000 |
| 12 | | PA | 17 | 5,350,864 | 0.318 | 0 | 1,321,415 | 0.000 | 0 | 417,395 | 0.000 | 12 | 2,646,305 | 0.453 | 0 | 741,611 | 0.000 |
| 13 | | HCSD | 0 | 5,350,864 | 0.000 | 0 | 1,321,415 | 0.000 | 0 | 417,395 | 0.000 | 0 | 2,646,305 | 0.000 | 0 | 741,611 | 0.000 |
| 14 | | BKT | 0 | 5,350,864 | 0.000 | 0 | 1,321,415 | 0.000 | 0 | 417,395 | 0.000 | 0 | 2,646,305 | 0.000 | 0 | 741,611 | 0.000 |
| 15 | Fatty Acid Oxidation Disorders | MCADD | 254 | 5,350,864 | 4.747 | 111 | 1,321,415 | 8.400 | 8 | 417,395 | 1.917 | 105 | 2,646,305 | 3.968 | 15 | 741,611 | 2.023 |
| 16 | | LCHADD | 9 | 5,350,864 | 0.168 | 0 | 1,321,415 | 0.000 | 0 | 417,395 | 0.000 | 0 | 2,646,305 | 0.000 | 0 | 741,611 | 0.000 |
| 17 | | VLCADD | 47 | 5,350,864 | 0.878 | 10 | 1,321,415 | 0.757 | 8 | 417,395 | 1.917 | 19 | 2,646,305 | 0.718 | 0 | 741,611 | 0.000 |
| 18 | | TFPD | 0 | 5,350,864 | 0.000 | 0 | 1,321,415 | 0.000 | 0 | 417,395 | 0.000 | 0 | 2,646,305 | 0.000 | 0 | 741,611 | 0.000 |
| 19 | | CDSP | 73 | 5,350,864 | 1.364 | 21 | 1,321,415 | 1.589 | 6 | 417,395 | 1.437 | 22 | 2,646,305 | 0.831 | 10 | 741,611 | 1.348 |
| 20 | Globinopathies | SCD+ | 922 | 5,350,864 | 17.231 | 8 | 1,321,415 | 0.605 | 809 | 417,395 | 193.821 | 72 | 2,646,305 | 2.721 | 0 | 741,611 | 0.000 |
| 21 | Endocrine Diseases | PCH | 2,672 | 5,350,864 | 49.936 | 510 | 1,321,415 | 38.595 | 71 | 417,395 | 17.010 | 1,595 | 2,646,305 | 60.273 | 404 | 741,611 | 54.476 |
| 22 | | CAH | 251 | 5,350,864 | 4.691 | 54 | 1,321,415 | 4.087 | 10 | 417,395 | 2.396 | 146 | 2,646,305 | 5.517 | 22 | 741,611 | 2.967 |
| 23 | Lysosomal Diseases | GSD-II | 13 | 643,218 | 2.021 | 0 | 158,845 | 0.000 | 0 | 50,174 | 0.000 | 6 | 310,287 | 1.934 | 0 | 89,147 | 0.000 |
| 24 | | MPS-I | 0 | 643,218 | 0.000 | 0 | 158,845 | 0.000 | 0 | 50,174 | 0.000 | 0 | 318,108 | 0.000 | 0 | 89,147 | 0.000 |
| 25 | Other | BIOT | 78 | 5,350,864 | 1.458 | 21 | 1,321,415 | 1.589 | 0 | 417,395 | 0.000 | 43 | 2,646,305 | 1.625 | 0 | 741,611 | 0.000 |
| 26 | | CF | 619 | 5,350,864 | 11.568 | 305 | 1,321,415 | 23.081 | 26 | 417,395 | 6.229 | 258 | 2,646,305 | 9.749 | 9 | 741,611 | 1.214 |
| 27 | | GALT | 59 | 5,350,864 | 1.103 | 26 | 1,321,415 | 1.968 | 0 | 417,395 | 0.000 | 23 | 2,646,305 | 0.869 | 0 | 741,611 | 0.000 |
| 28 | | SMA | 0 | 0 | 0.000 | 0 | 0 | 0.000 | 0 | 0 | 0.000 | 0 | 0 | 0.000 | 0 | 0 | 0.000 |

**Extended Dataset E2 [Part 3 of 3]**

| | Therapeutic Area | Disease Code | Aggregate | | Original Ancestry-Ethnicity Mapping | | | | | | | | Revised Ancestry-Ethnicity Mapping | | | | | | | |
|---|---|---|---|---|---|---|---|---|---|---|---|---|---|---|---|---|---|---|---|---|
| | | | gnomAD v4 P/LPs ≤ 1% | | gnomAD v4 P/LPs ≤ 1% | | gnomAD v4 P/LPs ≤ 1% | | gnomAD v4 P/LPs ≤ 1% | | gnomAD v4 P/LPs ≤ 1% | | gnomAD v4 P/LPs ≤ 1% | | gnomAD v4 P/LPs ≤ 1% | | gnomAD v4 P/LPs ≤ 1% | | gnomAD v4 P/LPs ≤ 1% | |
| | | | Aggregate | | White | | Black | | Admixed | | Pan Asian | | White | | Black | | Admixed | | Pan Asian | |
| | | | :100k | FD | :100k | FD | :100k | FD | :100k | FD | :100k | FD | :100k | FD | :100k | FD | :100k | FD | :100k | FD |
| 1 | | PKU | 8.581 | 2.07 | 11.780 | 1.56 | 0.697 | -3.78 | 3.843 | 1.43 | 0.384 | -8.07 | 10.745 | 1.42 | 0.697 | -3.78 | 3.843 | 1.43 | 0.384 | -8.07 |
| 2 | | MSUD | 0.070 | -5.31 | 0.155 | NA | 0.090 | NA | 0.020 | -26.90 | 0.007 | NA | 0.072 | NA | 0.090 | NA | 0.020 | -26.90 | 0.007 | NA |
| 3 | Amino Acid Disorders | HCU | 0.013 | NA | 0.014 | NA | 0.029 | NA | 0.042 | NA | 0.012 | NA | 0.015 | NA | 0.029 | NA | 0.042 | NA | 0.012 | NA |
| 4 | | CTLN-1 | 0.056 | -10.64 | 0.058 | NA | 0.036 | NA | 0.023 | -25.88 | 0.057 | NA | 0.063 | NA | 0.036 | NA | 0.023 | -25.88 | 0.057 | NA |
| 5 | | ASLD | 0.929 | 3.82 | 1.292 | NA | 0.135 | NA | 0.073 | NA | 0.010 | NA | 1.250 | NA | 0.135 | NA | 0.073 | NA | 0.010 | NA |
| 6 | | HT-1 | 0.107 | -1.57 | 0.148 | NA | 0.009 | NA | 0.015 | NA | 0.001 | NA | 0.110 | NA | 0.009 | NA | 0.015 | NA | 0.001 | NA |
| 7 | | IVA | 0.103 | -7.82 | 0.145 | -6.78 | 0.043 | NA | 0.022 | -31.26 | 0.006 | NA | 0.156 | -6.31 | 0.043 | NA | 0.022 | -31.26 | 0.006 | NA |
| 8 | | GA-1 | 0.262 | -3.56 | 0.297 | -3.31 | 0.450 | NA | 0.215 | -4.39 | 0.114 | NA | 0.313 | -3.14 | 0.450 | NA | 0.215 | -4.39 | 0.114 | NA |
| 9 | | HMGCLD | 0.002 | NA | 0.002 | NA | 0.002 | NA | 0.000 | NA | 0.000 | NA | 0.003 | NA | 0.002 | NA | 0.000 | NA | 0.000 | NA |
| 10 | Organic Acid Disorders | 3-MCC | 0.058 | -47.07 | 0.070 | -18.40 | 0.067 | -21.50 | 0.040 | -88.07 | 0.027 | -40.43 | 0.074 | -17.40 | 0.067 | -21.50 | 0.040 | -88.07 | 0.027 | -40.43 |
| 11 | | MMA | 0.080 | -9.81 | 0.092 | NA | 0.144 | NA | 0.012 | -60.58 | 0.460 | NA | 0.097 | NA | 0.144 | NA | 0.012 | -60.58 | 0.460 | NA |
| 12 | | PA | 0.009 | -34.02 | 0.007 | NA | 0.213 | NA | 0.004 | -110.49 | 0.017 | NA | 0.007 | NA | 0.213 | NA | 0.004 | -110.49 | 0.017 | NA |
| 13 | | HCSD | 0.001 | NA | 0.001 | NA | 0.000 | NA | 0.000 | NA | 0.031 | NA | 0.001 | NA | 0.000 | NA | 0.000 | NA | 0.031 | NA |
| 14 | | BKT | 0.004 | NA | 0.002 | NA | 0.000 | NA | 0.173 | NA | 0.011 | NA | 0.003 | NA | 0.000 | NA | 0.173 | NA | 0.011 | NA |
| 15 | | MCADD | 5.684 | 1.20 | 8.255 | -1.02 | 0.291 | -6.58 | 1.718 | -2.31 | 0.000 | -10,113.12 | 8.868 | 1.06 | 0.291 | -6.58 | 1.718 | -2.31 | 0.000 | -10,113.12 |
| 16 | Fatty Acid Oxidation Disorders | LCHADD | 0.204 | 1.22 | 0.326 | NA | 0.010 | NA | 0.078 | NA | 0.001 | NA | 0.250 | NA | 0.010 | NA | 0.078 | NA | 0.001 | NA |
| 17 | | VLCADD | 0.652 | -1.35 | 0.875 | 1.16 | 0.109 | -17.57 | 0.099 | -7.24 | 0.031 | NA | 0.932 | 1.23 | 0.109 | -17.57 | 0.099 | -7.24 | 0.031 | NA |
| 18 | | TFPD | 0.206 | NA | 0.327 | NA | 0.013 | NA | 0.080 | NA | 0.007 | NA | 0.251 | NA | 0.013 | NA | 0.080 | NA | 0.007 | NA |
| 19 | | CDSP | 0.239 | -5.70 | 0.289 | -5.50 | 0.031 | -45.82 | 0.087 | -9.55 | 1.483 | 1.10 | 0.309 | -5.14 | 0.031 | -45.82 | 0.087 | -9.55 | 1.483 | 1.10 |
| 20 | Globinopathies | SCD+ | 2.304 | -7.48 | 0.033 | -18.45 | 0.154 | -1,260.06 | 1.804 | -1.51 | 0.624 | NA | 0.035 | -17.08 | 0.154 | -1,260.06 | 1.804 | -1.51 | 0.624 | NA |
| 21 | Endocrine Diseases | PCH | 2.738 | -18.24 | 2.539 | -15.20 | 1.495 | -11.38 | 0.926 | -65.06 | 11.913 | -4.57 | 2.690 | -14.35 | 1.495 | -11.38 | 0.926 | -65.06 | 11.913 | -4.57 |
| 22 | | CAH | 22.760 | 4.85 | 22.703 | 5.56 | 10.948 | 4.57 | 5.657 | 1.03 | 1.702 | -1.74 | 24.224 | 5.93 | 10.948 | 4.57 | 5.657 | 1.03 | 1.702 | -1.74 |
| 23 | Lysosomal Diseases | GSD-II | 5.785 | 2.86 | 7.723 | NA | 1.874 | NA | 2.387 | 1.23 | 0.217 | NA | 8.251 | NA | 1.874 | NA | 2.387 | 1.23 | 0.217 | NA |
| 24 | | MPS-I | 0.423 | NA | 0.587 | NA | 0.018 | NA | 0.077 | NA | 0.000 | NA | 0.592 | NA | 0.018 | NA | 0.077 | NA | 0.000 | NA |
| 25 | | BIOT | 0.383 | -3.81 | 0.444 | -3.58 | 8.233 | NA | 0.196 | -8.30 | 0.013 | NA | 0.479 | -3.32 | 8.233 | NA | 0.196 | -8.30 | 0.013 | NA |
| 26 | Other | CF | 2.934 | -3.94 | 4.421 | -5.22 | 3.710 | -1.68 | 3.981 | -2.45 | 0.003 | -377.15 | 3.732 | -6.18 | 3.710 | -1.68 | 3.981 | -2.45 | 0.003 | -377.15 |
| 27 | | GALT | 0.934 | -1.18 | 1.196 | -1.65 | 1.726 | NA | 0.191 | -4.55 | 0.024 | NA | 1.290 | -1.53 | 1.726 | NA | 0.191 | -4.55 | 0.024 | NA |
| 28 | | SMA | 0.067 | NA | 0.006 | NA | 3.387 | NA | 0.154 | NA | 0.053 | NA | 0.000 | NA | 3.387 | NA | 0.154 | NA | 0.053 | NA |
